# Sub-Diffraction Stochastic Biosensing of Viruses in Untreated Plasma via Immuno-Janus Particle Agglutination and Flickering

**DOI:** 10.64898/2026.08.05.26359795

**Authors:** Tiger H. Shi, John A. Sinclair, Feng Gao, Satyajyoti Senapati, Tyler Moorman, Hsueh-Chia Chang

## Abstract

Viral diagnostics during early phases of infection are often limited by target scarcity and the deployment tempo. We significantly advance both quantitative accuracy and diagnostic throughput of viral agglutination assays with Immuno-Janus Particle (IJP) aggregation behavior that “flicker” stochastically with size-dependent statistics. By scrutinizing microscale blinking patterns of time series fluorescent videos, we decipher Brownian dynamics of individual IJP-Virus conjugates and IJP aggregates via windowed Itô stochastic analysis (termed the Culsans method). High-frequency rotational fluctuation is deconvolved from corrupting drifts caused by gravitational sedimentation and Brownian translational motion. This methodology enables a non-linear mapping of angular positions of detected IJPs and IJP aggregates to extract rotational diffusivity (*D_r_*) (and subsequently overall construct size) with superior linearity (*R*^2^ ≥ 0.85). The aggregation behavior exhibits a maximum when the IJP and viral particle concentrations are equal. The virion-bridged IJP-IJP conjugates significantly shift the detectable hydrodynamic diameter in the Poisson limit of reduced virus concentration with respect to IJPs, pushing the limit of detection (LOD) to 10^3^ - 10^4^ virions per mL in untreated human plasma. This tunable platform offers a rapid, low-volume, and scalable alternative to lab-based RT-PCR, bridging the gap between virion sensitivity and field-readiness.

## 1 Introduction

Prompt diagnosis early into disease onset is critical for improved patient and community health outcomes during viral outbreaks. The technologies necessary for prophylactic strategies are constantly at odds between target sensitivity and deployment tempo. During outbreak scenarios such as during that of SARS CoV 2, patients exhibiting low viral burdens are primarily early-stage infections.^1^ During this phase, treatment is still relatively easy, viral spread is manageable, and hospital burden may be reduced.^2,3^ Yet, this target scarcity creates an imposing bottleneck for clinical intervention in these highly relevant asymptomatic stages of infection during which patients still exhibit high transmissibility.^2,4^

During early stages of infection, viral load in biosamples ranges between 10^2^ to 10^5^ copies/mL in blood and mucus.^3,5^ In the case of SARS CoV 2 pandemic, patients exhibiting high (10^8^ copies/mL), medium (10^5^ copies/mL), and low (10^2^ copies/mL) were shown to be equally distributed in the patient population.^1^ The standard for a high-tempo, population scale test is the lateral flow assay (LFA), which is prized for its readiness for mass manufacturing, mass distribution, and rapid at-home screening. Yet, more than 70% of infected individuals present viral burdens undetectable by standard LFAs at a limit of detection (LOD) of 10^6^ virions/mL.^6^ The deployment of high-sensitivity Reverse Transcription Polymerase Chain Reaction (RT-PCR) assays with an LOD of 10^3^ virions/mL in the original untreated sample is unfeasible due to its cost in scalability due to equipment, facility, and personnel requirements stipulated by its requisite pretreatment steps.^6^ Still, nearly 15% of the effected patient population slip through.^1^

Agglutination-based assays are a prime class of candidate platforms for rapid, cost-effective, and field-deployable virus detection.^7,8^ These platforms leverage colloidal or cell-based aggregation driven by multivalent virus binding. However, visual agglutination assays typically suffer from poor quantitative output, with detection limits at or above 10^6^ virions/mL.^9,10^ Bulk optical techniques, e.g. spectrophotometry and dynamic light scattering (DLS), improve quantification capability by measuring aggregate size distributions, yet still suffer from limitations in the optical noise floor and strong Rayleigh scattering bias 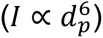 drowning out signal from smaller particles of interest.^11^ Consequently, in complex, unpurified, physiological media containing dilute (< 10^6^ aggregates/mL) aggregate populations, label-free light scattering techniques struggle to track individual target aggregates with reasonable accuracy due to high background scattering from medium components limiting the reliability of particle sizing via the Einstein-Stokes diffusivity relationship.

We have previously reported an Immuno-Janus Particle (IJP) technology^12,13^ that can capture the Brownian rotational dynamics of a sub-diffraction particle (500 nm to 1 μm) even when tested in a complex, biological medium. These particles are composed of a fluorescent polystyrene (PS) hemisphere and a dark antibody-functionalized gold hemisphere. When recorded over time, the rotational diffusion causes these particles to “flicker” as the camera observes varying amounts of the fluorescent and gold hemispheres. As demonstrated in our prior works, fluorescent flickering behaviors of individual IJPs are clearly detectable in a complex physiological fluid.

The presence and shape of extracellular vesicle (EV) and viral targets are indiscernible from within the image due to their sizes (50 nm – 200 nm) being well below the diffraction limit of light microscopy (∼300 nm). These nano-scale vesicular entities are still observable by their effects on the flickering behavior of the IJPs hosting them due to the Einstein-Stokes-Debye rotational diffusivity *D_r_* scaling as d_p_^-3^, where d_p_ is the effective rotational hydrodynamic diameter of a spherical particle. (The concurrent size-scaled Einstein-Stokes translational diffusivity *D_t_* scaling with d_p_^-1^ necessitates that flickering due to translational activity must be filtered for accurate estimation of *D_r_*.) The blinking signal is sensitively affected by changes in IJP hydrodynamic diameter via the specific docking of nanocarriers larger than 30 nm in diameter, such as extracellular vesicles (EVs).^12,13^ This size-based gating effect also renders the IJP platform to be insensitive to interference by non-specifically adherent proteins (< 30 nm) allowing for robust detection in complex biofluids like untreated plasma. As viruses are within the same size range and also exhibit characteristic surface markers such as the spike glycoprotein in SARS CoV 2, the IJP is a strong candidate for a rapid, low-volume test for early viral infections. In this work, we significantly enhance the sensitivity of this established technique by extending to an IJP agglutination assay.

The previous implementation of the IJP analysis is only capable of achieving LODs no smaller than 10^6^ to 10^7^ particles/mL, which is comparable to an LFA and is several orders of magnitude greater than the necessary LOD for practical use in detecting early stage viral loads.^12^ The first bottleneck lies with the data analysis pipeline which relies on a rough approximation of non-linear, non-periodic behavior using a Continuous Wavelet Transform (CWT) which is designed for interpreting periodic behaviors. The biosensor’s LOD is limited by approximation using only population behavior. Another limitation of the current implementation of IJPs lies in that the observation of particle blinking changes caused by virus binding events is often corrupted by the influence of ballistic gravitational sedimentation and translational Brownian dynamics through the image volume. The final bottleneck is a rigorous analysis of multi-IJP aggregation behavior, rather than only characterizing IJP-Virus conjugates. By analyzing virus-bridged IJP aggregation events, we overcome all these obstacles to realize a pretreatment-free, field-deployable, and quantitative agglutination assay for viruses exhibiting target sensitivity that rivals RT-PCR platforms.

This work develops the precise stochastic physics framework governing the rotational diffusion of individual Immuno-Janus Particles (IJPs) and further extending it to IJP aggregates. We demonstrate that *D_r_* is most faithfully estimated via a windowed Itô stochastic analysis, which we name the Culsans method. This methodology leverages single-particle statistics allowing us to interrogate individual binding events and push the LOD down to 10^3^ particles/mL the range of practical deployment. We show that IJP concentration can be optimized with respect to viral concentrations to actively shift the dynamic range of the sensor to operate within a wide range of concentration contexts. Finally, we report an IJP aggregation mechanism at the Poisson limit that explains the significant increase in sensitivity of this noise-sensing platform.

Through rigorous computational modeling of these microscale physical dynamics, we break free of the traditional semi-empirical sensor configurations to achieve Poisson-limited single-particle sensitivity and uncover a novel aggregation-enhanced signal amplification mechanism tailored for early-stage, ultra-sensitive pathogen detection.

## 2 Results

### 2.1 Intensity fluctuation by one-dimensional Brownian rotation in IJPs

Without action caused by deterministic sedimentation or translational Brownian motion, environmental thermal kicks will apply statistically random torque varying in size and direction. Brownian rotational diffusion (*D_r_*) operates analogously to Brownian translational diffusion (*D_t_*) (Supplementary Figure S3). For a given time step *δt*, rather than experience statistically random translational steps defined by *D_t_*_,*i*_ along each independent spatial axis *i*, the particle experiences statistically random rotational steps defined by *D_r_*_,*j*_ about each independent rotational axis *j*. The recognition that this rotational behavior causes observable fluctuations in the detected fluorescent intensity *I*(*t*) for each IJP underpins the core of our theory. To ensure thoroughness, we specify the geometry and degrees of freedom available. The changes in local brightness do not directly correlate to angular deviations in 3D spherical space, but rather these fluctuations are a proxy for the exposed fluorescent area in a 2D projection (Δ*I* = *I*(*t*_1_) − *I*(*t*_0_)).

We define each movement as a relative rotation between two vectors (Figure 1): (1) *n̂* corresponding to the fixed vector originating from the center of the IJP pointing orthogonally to an observer away from the image plane, and (2) *m̂* corresponding to the free spinning vector in ℝ^3^-space originating from the center of the IJP and traversing the fluorescent hemisphere in a direction perpendicular to the dividing plane of the two hemispheres. The particle exhibits two rotational degrees of freedom (*θ*, *ϕ*) in spherical geometry and displays an auxiliary rotational degree of freedom (*ψ*) about the vector *m̂*. Due to rotational symmetry about *m̂* and *n̂*, rotations in *ψ* and *ϕ*, respectively, do not alter the detected fluorescence intensity, i.e. the total fluorescent area observable by the viewer. This renders only one relevant rotational degree of freedom *θ*. We demonstrate the relationship Δ*I* = *f*(Δ*θ*), i.e. intensity variation is only a function of *θ*, by injecting thermal kicks upon a simulated particle along two linearly independent rotational axes that are not the absolute *θ*, *ϕ*, nor *ψ* within the simulated spherical space.

**Figure 1.**
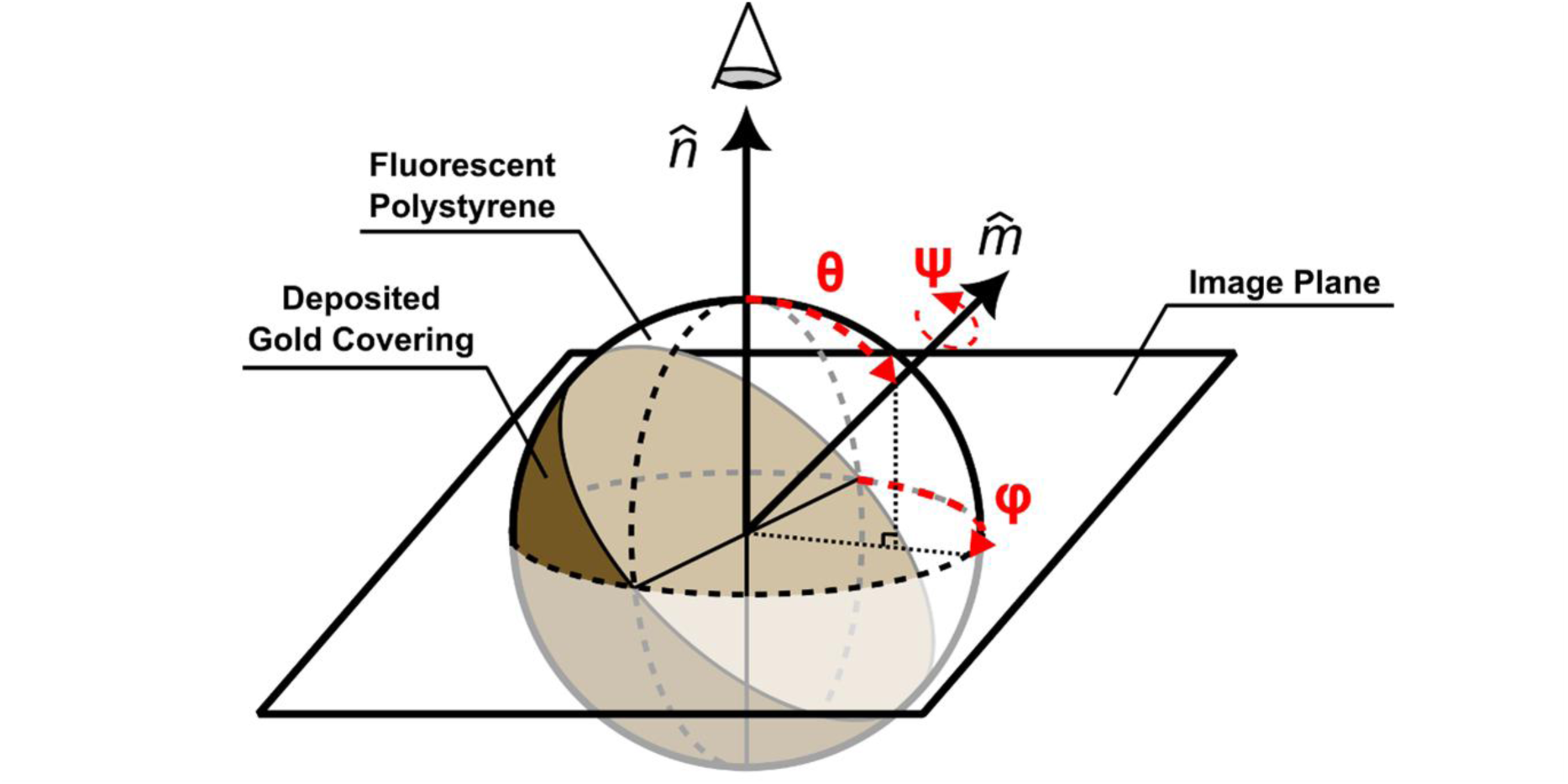
Immuno-Janus Particle (IJP) in spherical coordinates as observed orthogonally to and impinged within an image plane. The rotational modes are represented by three degrees of freedom (red). Characteristic fixed *n̂* and free *m̂* unit vectors.

To demonstrate the decomposability of these two rotational moments, we simulated translationally static blinking IJPs with simulation rate of 300 steps per second and capture frame rate of 10 frames per second (Figure 2a). Simulated rotational steps were independently calculated for both (*θ*, *ϕ*) components in absolute coordinates using the Einstein-Stokes equation for rotational diffusivity. The simulation space was set up so that the camera position vector *n̂* is (*θ*, *ϕ*) = (*π*⁄3, − *π*⁄3), or 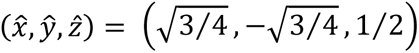 (Figure 2b), allowing for the demonstration of the decoupling of two angular degrees of freedom given an arbitrary observation reference point. Setting *n̂* as the new reference axis for system renormalization, we may transform the position vector *m̂* from any given position in absolute spherical coordinate space to a new auxiliary position space. This is achieved by projecting *m̂* onto the plane orthogonal to *n̂* giving the auxiliary coordinates:

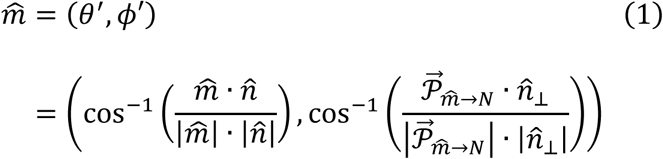

where *θ*^′^ and *ϕ*^′^ are transformed spherical angular coordinates with respect to *n̂*, *N* is the plane orthogonal to *n̂* and intersecting with the origin, *n̂*_⊥_ is an arbitrary reference vector 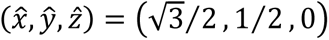 within *N* used to determine the *ϕ*^′^ component, and 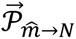 is the projection of *m̂* onto *N* determined as 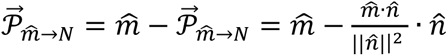. Due to rotational symmetry about *n̂*, fluctuation in observed intensity Δ*I* is correlated with |*θ*^′^| rather than with *θ*.

**Figure 2.**
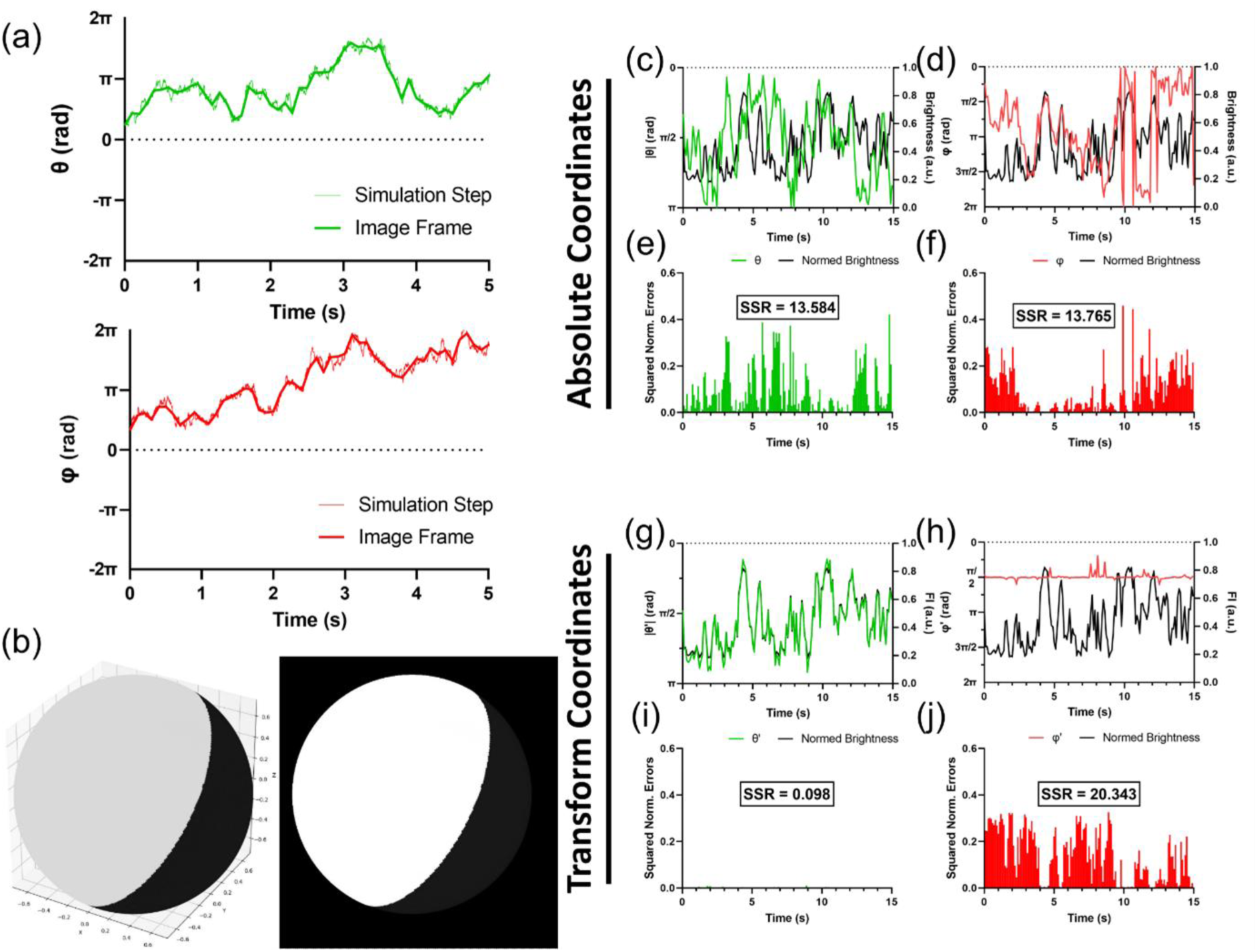
Angular component isolation demonstrates rotational parameter space to be composed of two mutually independent, spherically orthogonal degrees of freedom that can be isolated by defining coordinates based on arbitrary viewing angle. (a) Absolute angular coordinates of a simulated particle over 5 seconds. The black lines show the angular components recorded based on frame rate. The red lines show the angular components recorded based on simulation rate. The top figure is *θ* ∈ [−*π*, *π*] and the bottom figure is *ϕ* ∈ [−2*π*, 2*π*]. (b) Representative images of the observable side of two simulated particles, where the left image shows the coordinates and the right image shows the final image. (c-f) Comparison of absolute coordinates (*θ*, *ϕ*) to the measured brightness values of each frame over the course of 15 seconds. (c, d) shows (|*θ*|, *ϕ*), respectively, each plotted against brightness. (e, f) shows the squared, normalized errors of (*θ*, *ϕ*) against brightness. (g-i) Comparison of absolute coordinates (|*θ*^′^|, *ϕ*^′^) to the measured brightness values of each frame over the course of 15 seconds. (g, h) shows (*θ*^′^, *ϕ*^′^), respectively, each plotted against brightness. (i, j) shows the squared, normalized errors of (*θ*^′^, *ϕ*^′^) against brightness.

When observing the absolute coordinates (Figure 2c-f), we notice that the fluctuations in |*θ*′| and *ϕ* values correlate poorly with the observed brightness values for each frame. However, when converted to the transformed coordinates with respect to the viewport (Figure 2g-j), the |*θ*′| component almost perfectly captures the observed blinking phenomenon (Figure 2g, i), while the *ϕ*′ becomes entirely uncorrelated (Figure 2h, j). Error in the *θ*′ estimate only arises as |*θ*′| → 0 or |*θ*′| → *π* and can be attributed to the estimation limit of the non-linear mapping between normalized brightness levels and angular orientation (discussed further in Section 2.3). These observations allow us to conclude that: (1) any given rotation in spherical geometry can be decoupled into any two rotationally orthogonal degrees of freedom based on an arbitrarily defined reference axis *n̂*, (2) the ability to decouple rotational degrees of freedom is not an artifact of the simulation conditions, and (3) fluctuations in observed brightness level of a static IJP given an arbitrary observation axis can be fully attributed to changes in the decoupled rotation in the redefined *θ*′ and fully independent of the new *ϕ*′.

With the key understanding that Δ*I* = *f*(Δ*θ*), we may confine and simplify the analysis to rotational dimension *θ* by modeling the relevant rotational diffusive steps *dθ*(*t*) as a purely stochastic behavior independent of translational diffusion. Doing so, we yield a Gaussian distribution of steps to model changes in *θ*:

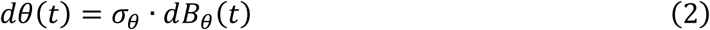

where *dB_θ_* is a Wiener (Brownian motion) process with each process step following a normal distribution 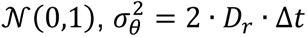 is the timestep-dependent spread of *dθ* with respect to *dB_θ_*, and 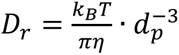 is the rotational diffusivity of the IJP. Taken together, this formal definition – with the correlation between the observed brightness of a static particle and its orientation about the (*n̂* × *m̂*)-axis defined as *θ*^′^ – allows us to describe the relationship between the transient brightness variations caused solely by rotational contributions *I_θ_* and the effective IJP diameter *d_p_*.

In the observable hemisphere, all viewable conformations can be directly characterized by the angle cos *θ* = *n̂* · *m̂*, where "Maximum Fluorescence" corresponds to |*θ*| = 0, "Half Fluorescence" corresponds to |*θ*| = *π*⁄2, and "Minimum Fluorescence" corresponds to |*θ*| = *π*. Measuring orthogonally to the axis of rotation within the image plane, the length of fractional fluorescence along the observed diameter of the projection can be expressed as 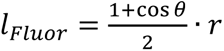. Solving for the area of the fractional fluorescent projection, we yield 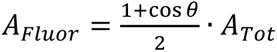, where *A_Tot_* is the total area of the circular projection of the spherical particle 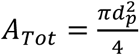. If we assume that fluorescent intensity *I_θ_* is directly related to the proportional exposed fluorescent area *A_Fluor_*⁄*A_Tot_*, we can set:

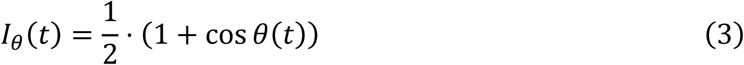

to be the bound normalized fluorescent intensity caused by rotational diffusion. Notice that this function yields a maximum bound of *I_θ_* = 1 when the directional vector is parallel with the viewing axis *θ* = 0, and a minimum bound of *I_θ_* = 0 when the directional vector is antiparallel with the viewing axis *θ* = ±*π*.

Solving using the Itô Lemma, we achieve the statistical components of the brightness factors:

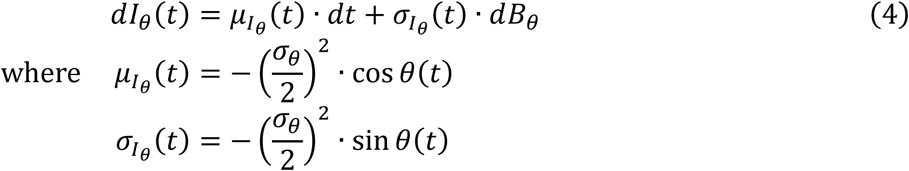

Note that the deterministic component drifts with respect to 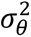 while the stochastic component simply inherits the *σ_θ_* scaling per the prior definition of *dθ*.

### 2.2 Signal amplitude normalization to correct translational ballistic and diffusive drifts

Comparing fluorescent intensity evolutions of simulated particles experiencing only rotational degrees of freedom (Figure 2 and Figure 3c) against observed fluorescent tracks of real IJPs (Figure 3b), we notice additional base-line drifts superimposed upon the base fluorescent evolution. These unaccounted degrees of freedom exhibit characteristic periods distinctly longer than those of rotational fluctuations. These slow drifts – relative to the action caused by rotation – are caused by the motion of the IJP in and out of the focal plane (Figure 3a) due to gravitational settling and translational diffusion normal to the focal plane. We hence developed a strategy for deconvolving the slow ballistic sedimentation and orthogonal translational diffusion dynamics via a workflow involving windowed averaging and renormalization.

**Figure 3.**
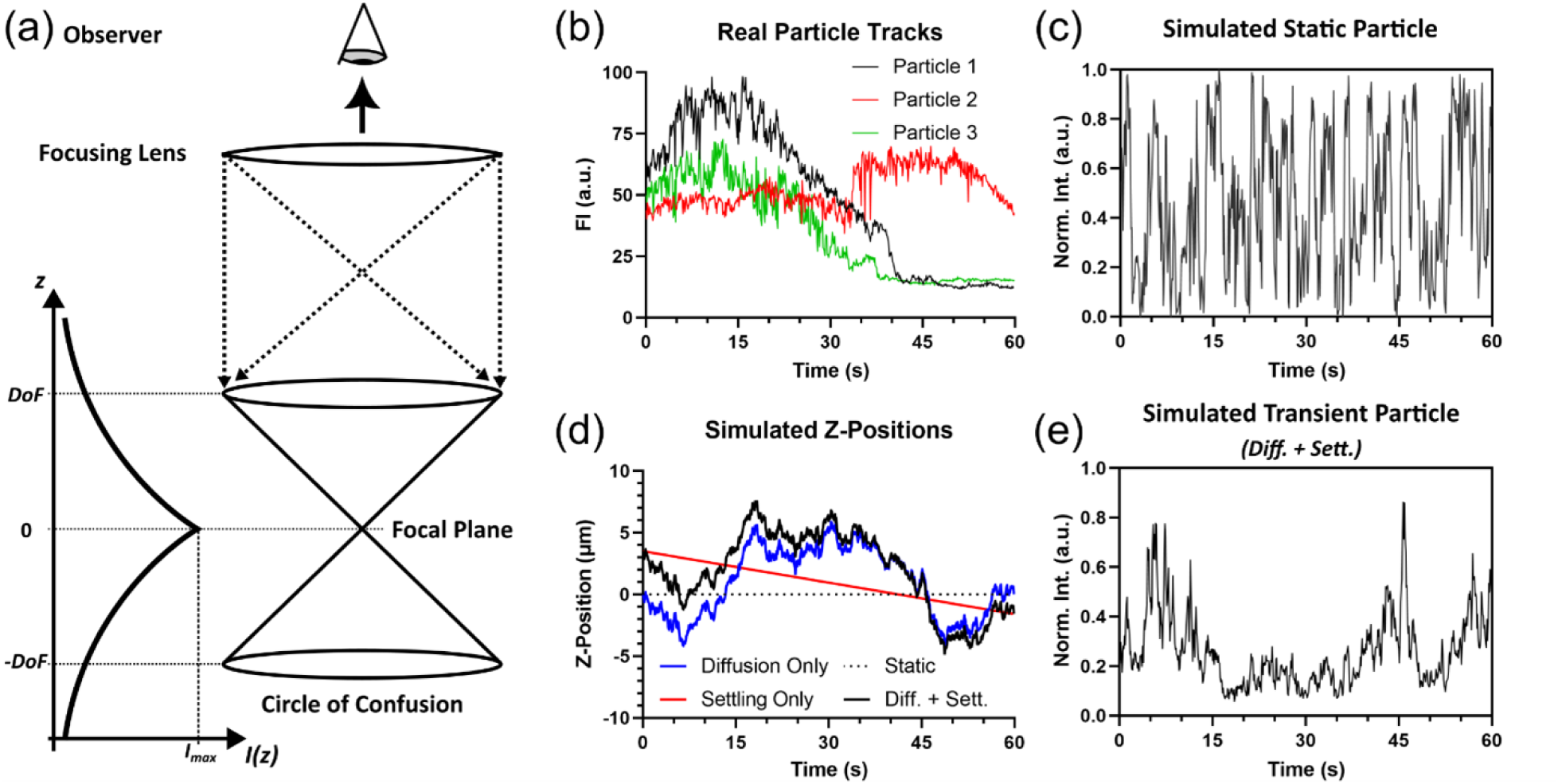
*Z*-axis translational behaviors of rotating particles convolute two mechanisms of fluorescence modulation. (a)Diagram illustrating the differences in intensity of images observed through a lens depending on the object’s position in relation to the focal plane. Depth of field (*DoF*) in this work represents a distance from the focal plane such that the resultant circle of confusion for a given particle has twice the observed area than when it was in focus. (b) Select brightness time series tracks over the course of one minute of three independent 1 *μm* IJPs suspended in 1× PBS without incubation with any capture targets. (c) Brightness time series track over the course of one minute of a simulated static 1 *μm* IJP without translational motion. (d) Simulated positions of 1 *μm* IJP in the *Z*-axial direction when static (dotted), under the influence of only Brownian translational diffusion (blue), under the influence of only gravitational settling (red), and combined diffusive and gravitational forcing (black). (e) Brightness time series track of the same 1 *μm* IJP from (c) while subjected to combined dynamic diffusive and gravitational forcing in the *Z*-axial direction through the focal plane of the simulated image plane.

Gravitational settling is a purely deterministic behavior given as:

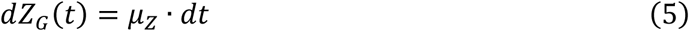

where the settling velocity 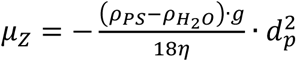 is the resultant terminal rate of the particle with diameter *d_p_* accounting for buoyancy Δ*ρ_i_*_,*j*_ and drag forces *η*. We can assume that all particles within frame have achieved terminal velocity due to the timescale of the experimental procedure leading up to measurement (several seconds to a minute) being longer than the requisite acceleration time required to achieve terminal velocity (< 1 µs). All sample parameters, such as viscosity and density, may be assumed to be those of water at room temperature, as the sample is procedurally diluted 10 × in 1X PBS causing minimal variation due to patient-to-patient differences.

Translational diffusion along the *Z*-axis normal to the viewing plane is a purely stochastic behavior given as:

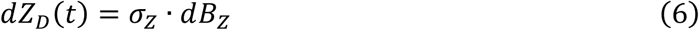

where *B_Z_* is a Wiener process (Brownian motion) independent of *B_θ_* – the process linked to rotational perturbations, 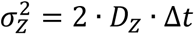 is the timestep-dependent spread of *dZ_D_* with respect to *dB_Z_*, and 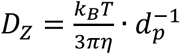 is the translational diffusivity of the particle with inverse correlation with particle size. Likewise with ballistic settling, All sample parameters, such as viscosity and density, may be assumed to be those of water at room temperature. As both the deterministic and stochastic components impact the *Z*-axis trajectory of the particle, their contributions can be superimposed into a singular translational Itô Process featuring both a deterministic and stochastic component. This term encompasses the position change in the *Z*-axial direction due to both ballistic sedimentation and diffusive translation:

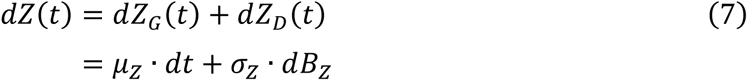

Compiling the contribution of the particle’s translational motion to the final fluorescence time series, we assume fluorescence signal decays per the Inverse Square Law of Radiation. We define this correctional contribution by IJP translation into fluorescent intensity as:

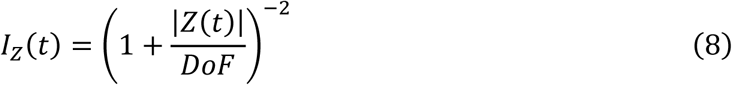

where the signal is normalized to the maximum intensity achieved when the particle is in the focal plane (*z* = 0). Furthermore, any position above or below the focal plane (*z* > 0) ∨ (*z* < 0) leads to an inverse square reduction of detected signal. We define an arbitrary Depth of Field (*DoF*) length scale that determines the distance from the focal plane the object needs to be in order to yield half the maximum intensity. This value is variable for each experiment set and accounts for variations in equipment and environmental conditions. In our analysis, we never need to calculate such a value. Our simulation work uses a variety of predefined *DoF*-values across every simulated particle track to test robustness of our developed algorithm. Experimentally, the algorithm operates only using the normalized quantity 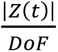 to define a weighting term which is internally normalized based on all other particle tracks within a given video capture. From the results of both simulations and experiments using our custom algorithm (discussed in Section 2.3), we find that the workflow is agnostic to the actual length scale value of the *DoF*.

As *I_Z_*(*t*) is related to distance from the focal plane |*Z*(*t*)| rather than the exact position *Z*(*t*), we declare the differential *d*|*Z*(*t*)| is the same as *dZ*(*t*) for all *Z*(*t*) > 0. When *Z*(*t*) < 0, the values are negated. Applying the Itô Lemma, we find that the step variations of this intensity time series *dI_Z_*(*t*) may be expressed as itself an Itô process with respect to *t* and *B_Z_*:

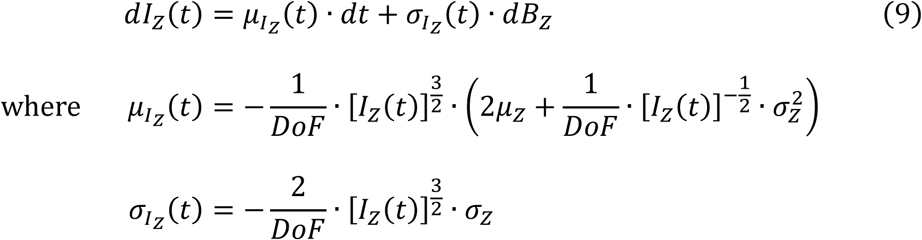

Note that the deterministic component of the brightness changes is linked to both *μ_Z_* and 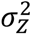, while the stochastic component remains solely influenced by *σ_Z_* · *dB_Z_*.

With intensity components due to both the paired ballistic-diffusive influences on the vertical translational dynamics *I_Z_*(*t*) and the diffusive influence on the rotational dynamics *I_θ_*(*t*) established, we evaluate their composite form, i.e. the true observable fluorescence time series *I*(*t*). The term *I_Z_*(*t*) represents the local maximum intensity achievable for a given particle where the entire fluorescent hemisphere is directed at the observer. We define *I_Z_*(*t*) as the upper envelope bounding *I_θ_*(*t*), such that each *I*(*t_i_*) intensity step is the associated *I_θ_*(*t_i_*) intensity step for a non-translating IJP scaled to the local maximum defined by *I_Z_*(*t_i_*). However, we empirically find that the gold-plated hemisphere with the fluorescence obscured is not perfectly dark, in that local intensity fluctuations do not reach zero, i.e. *I*(*t_i_*) ≠ 0, unless the regional intensity is already near zero, i.e. 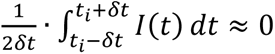 for *δt* values much larger than the video capture frame rate but much smaller than the total duration of the video. To account for this, we assume a minimum intensity of the darkened particle proportional to its maximum intensity min(*I*(*t*)) = *α* · *I_Z_*(*t*), where the proportionality parameter *α* ∈ [0,1] is fitted empirically for each IJP fluorescence intensity time series. With the above relationships, we define the expected observed fluorescent intensity time series and its differential as an Itô process:

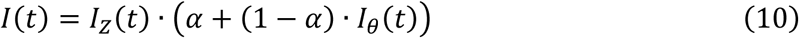

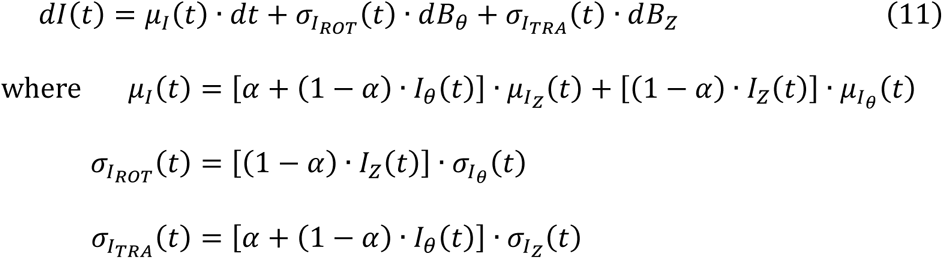

By simulating the particle to experience both rotational and translational degrees of freedom, we find that the predicted time series (Figure 3e) more closely resembles the observed time series (Figure 3b) than does the simulated time series only involving rotation (Figure 3c). In the differential form of the composite intensity via Eq. (11), the determinative component *μ_I_*(*t*) demonstrates the convolution between those of the two independent processes *I_θ_*(*t*) and *I_Z_*(*t*). Furthermore, notice that the differential contains direct and independent contributions from both of the Brownian diffusive processes *B_θ_* and *B_Z_* scaled by 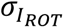 and 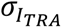, respectively.

### 2.3 Signal deconvolution via the Culsans method

With the underlying mechanism established, we developed a deconvolution procedure given an arbitrary total intensity time series. The methodology follows generally three main steps (Figure 4): (1) optimized time-averaged anchoring to approximate *I_Z_*(*t*) and *Z*(*t*), (2) envelope bounding and signal dilation by *α*-fitting to approximate *I_θ_*(*t*), and (3) non-linear mapping of *I_θ_*(*t*) and *dI_θ_*(*t*) to approximate the full distribution of *θ*(*t*) within a defined time window.

**Figure 4.**
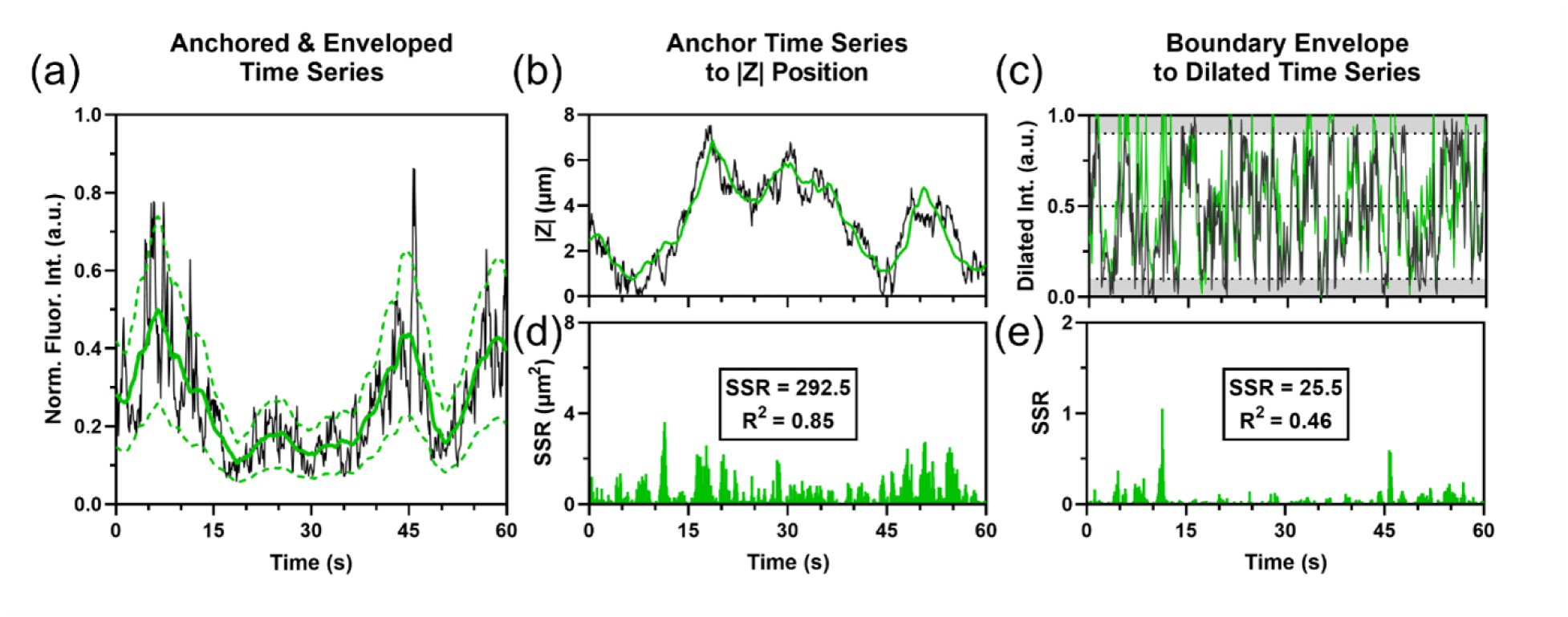
Representative workflow for deconvolving short-timescale rotational brightness dynamics from long-timescale translational brightness dynamics for any arbitrary dynamic time series. Sample time series is from a simulated 1000 nm IJP with *α* = 0.30 and *DoF* = 4500 nm. (a) Given a normalized raw fluorescent intensity time series, the fluctuating central tendency anchor line (solid green line) is established through time averaging of the raw series. Upper and lower bound time series (dotted green lines) are determined through vertical scaling of the anchor line by a parameter *α*. In this case, averaging time shown is Δ*t_W_* = 5.0 seconds the fitted fractional brightness was *α* = 0.2270. (b) Central tendency can be isolated mapped directly to approximate the distance to the focal plane via Eq. (11). (c) The region between the upper and lower intensity boundaries is dilated so that the upper bound is set to 1.0 and the lower boundary is set to 0.0. Doing so, the brightness time evolution of the particle approximates its static equivalent, where intensity variations can be solely attributed to rotational fluctuations. Based on this approximation, the particle size can be estimated using the Culsans Method. (d-e) Sum squared residuals and *R*^2^-values for isolation of (d) translational and (e) rotational components.

#### 2.3.1 Time-scale optimization and the isolation of I_Z_(t) and I_θ_(t) from the measured I(t)

We understand that the relevant time period for the translational degree of freedom is longer than that of the rotational degree of freedom. As such, we employ a strategy of time averaging to establish an estimated long-time fluctuation dynamic that closely approximates the contributions of the translational motion of the particle. We use Eq. (12) to generate a moving time-averaged anchor series that traces the movement of the central tendency of the fluorescence series. The time series *I_Mid_*(*t*) is the discrete time-averaged evolution which traces the central trend of *I*(*t*) time series via the specified averaging window kernel Δ*t_W_*.

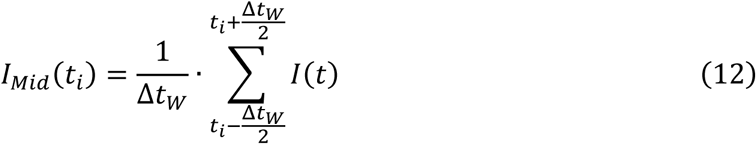

This strategy operates on a key assumption. During the time it takes for any significantly measurable changes in signal intensity caused by translation, rotational fluctuations would have covered a large enough range of potential states. In-so-far that, the average within the time window would yield the signal intensity attributable to approximately the midpoint between the brightest and dimmest rotational states.

There are two fitting parameters to our statistical analysis: (1) the averaging window Δ*t_W_* and (2) the fractional brightness *α* that defines the bounding of *I_θ_*(*t*) about the central trend *I_Mid_*(*t*). The parameter *α* ∈ (0,1) is determined empirically for each time series. Due to the differences in degree of separation between the rotational and translational time scales for particles of differing length scales, the averaging window Δ*t_W_* is optimized to a set scalar quantity over a large parameter space. We determine this optimal window size for practical IJP dimensions (500 – 5000 nm in diameter). By dilating a zone about *I_Mid_*(*t*) using Eq. (13) and (14), we can extract the upper bound *I_Fluor_*(*t*) and lower bound *I_Gold_*(*t*) envelopes, where *α* is the fractional brightness of the Gold hemisphere of the IJP in relation to the Fluorescent hemisphere (*α* = *I_Gold_*⁄*I_Fluor_*).

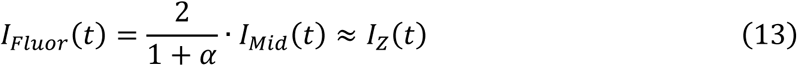

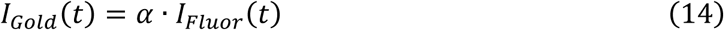

In Supplementary Figure S1a, we show that under translationally dynamic conditions the fitting algorithm recaptures the true *α*-values for medium and large size particles quite robustly. Additionally, we find that in our experiments the *α* of our IJPs regardless of IJP or virus concentration is typically stable at *α* = 0.45 ± 0.15 across 7850 independent simulated particle tracks.

Using the upper bound *I_Fluor_*(*t*) and the identity used for simulation in Eq. (8), we approximate *I_Fluor_*(*t*) to be *I_Z_*(*t*) and yield the estimated distance between the IJP and the focal plane:

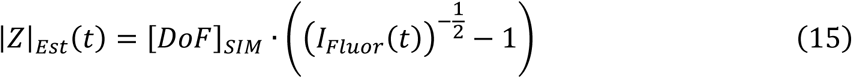

Using both the upper *I_Fluor_*(*t*) and lower *I_Gold_*(*t*) bounds, we can dilate intensity time series using Eq. (16) and isolate the short-time period fluctuations from the long-time period evolution. This operation recontextualizes the raw brightness time series {*I*(*t*) | *I*(*t*) ∈ [*I_Gold_*(*t*), *I_Fluor_*(*t*)]} as the range normalized approximate rotational brightness contribution {*I_θ_*(*t*) | *I_θ_*(*t*) ∈ [0,1]}.

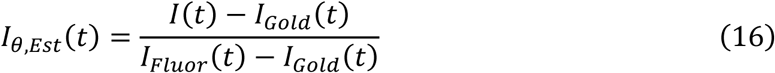

To gauge the accuracy of the position approximation, we coined the Sum Squared Residual (*SSR*) value as the Movement Isolation Error Rate (*ζ*) for a given particle time series. This value is generated by the comparison of the |*Z*|*_Est_*(*t*) value calculated to the ground truth simulated path |*Z*|*_SIM_*(*t*). Note that we are comparing the inverse of the displacements as the fluorescence is more sensitive to changes closer to the focal plane |*Z*|(*t*) ≪ 1 than to those further away |*Z*|(*t*) ≫ 0.

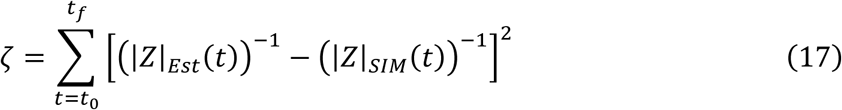

For the accuracy of the dilation operation, we similarly coined the *SSR*-value referred to as the Signal Recovery Error Rate (*ρ*). This quantity compares the approximated static-particle time series *I_θ_*_,*Est*_(*t*) to the ground truth fluorescence intensity of a simulated static-particle *I_θ_*_,*SIM*_(*t*) subject to the exact same rotational changes as the dynamic particle but without the translational component.

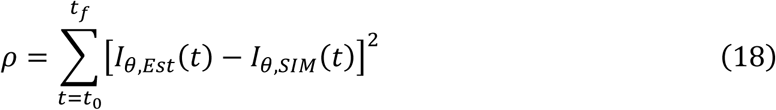

To simplify optimization, the two error rates were combined into a singular Convoluted Error Rate (*SSR_Conv_*) which was used as an objective function to be minimized.

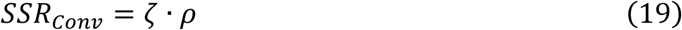

We simulated 24 individual particles for each of 12 unique diameter size bins ranging from 500 to 5000 nm. Each particle is affected individually by randomly generated rotational and translational impulses based on their assigned size bins (akin to methodology found in Sections 2.1 and 2.2). Thereafter, we applied the above normalization methodology to find the window kernel Δ*t_W_* that is most suitable given the relevant size bin range.

In Figure 5, we see the characteristics of the above approximation approach. If averaging times are too low or too high, the convoluted error grows suggesting poor isolation of translational and rotational effects. Additionally, this threshold is variable based on particle size as well. With particles of diameters within our IJP operating range (1250 - 3000 nm), the optimal averaging period is higher than that of smaller and larger particles. This size-based difference in effective approximation is due to the differences between three key characteristic times: (1) the amount of time required for *Z*-dynamics (diffusive/ballistic) to make a detectable in brightness levels, (2) the amount of time required for rotational motion to make a detectable change in brightness levels, and (3) the selected averaging time threshold that separates these two values.

**Figure 5.**
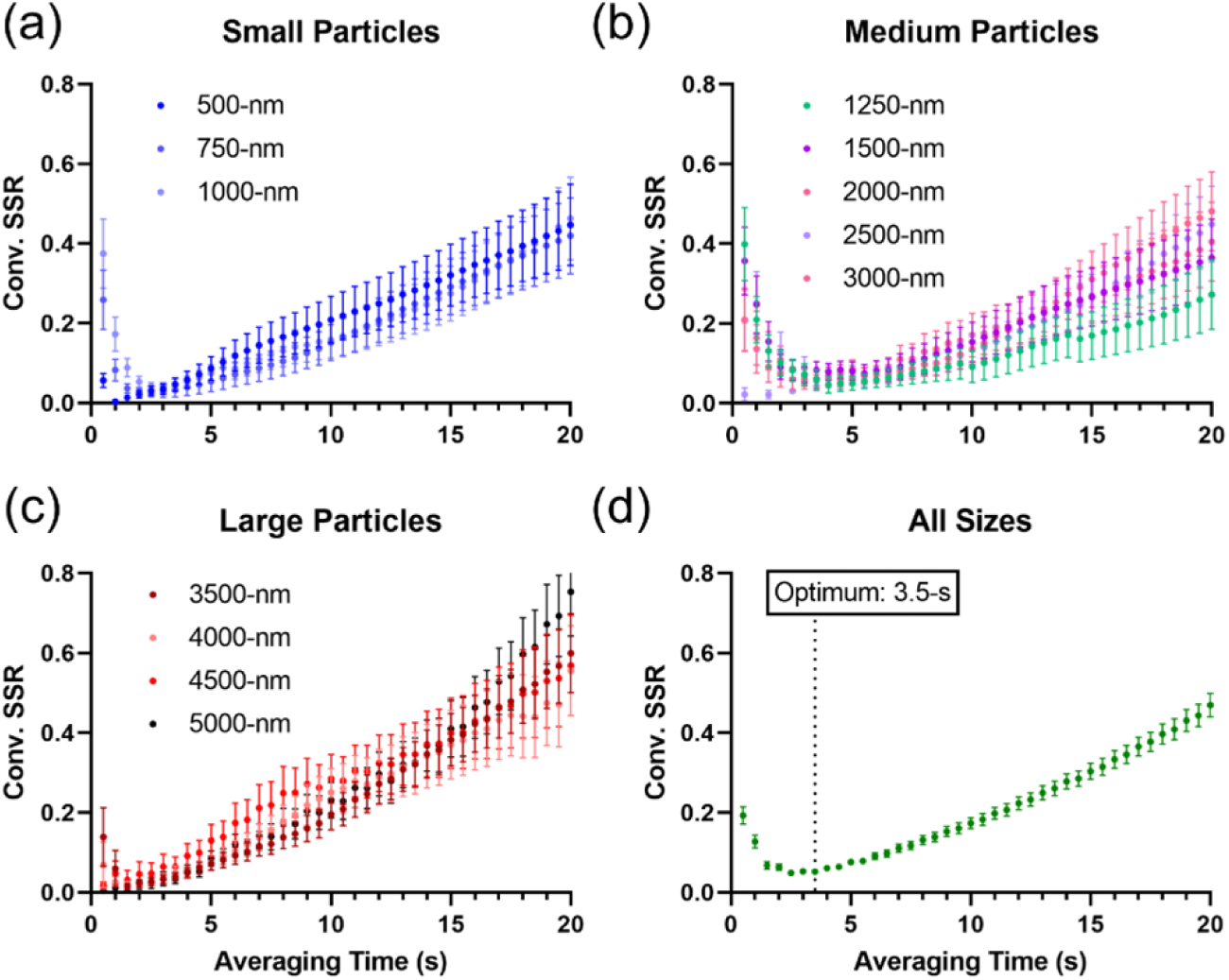
Grid search across averaging window lengths for time series of particles of various sizes (n = 24 per size) where the *SSR_Conv_* is calculated by Eq. (19). (a - c) Grid search results for particles of (a) small (500 - 1250 nm), (b) medium (1500 - 3000 nm), and (c) large (3500 - 5000 nm) diameters. (d) Aggregated grid search plot for all particles from 500 to 5000 nm. The averaging time window that yielded the overall lowest convolved error is indicated to be 3.5 seconds. All error bars across all plots represent the standard error of mean.

For the case of smaller particles, both the relevant translational and rotational time scales are shorter, due to their respective diffusivities scaling by 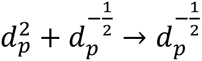 and 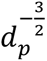. This means that a feasible averaging period that thresholds between the two competing time scales will be small in magnitude and exist within a narrow band of values. For large particles, the scaling via 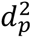 of ballistic gravitational settling dominates so significantly that the particle barely remains in frame for the length of a video. This leads to the majority of the time series appearing dark as the sensor is not capable of observing the particle while it is out of the camera’s focal volume. Time averaging using longer time periods leads to the counterintuitive erasing the brief period of time where the particle is in frame. However, for the key case of mid-sized particles on the scale of 1 - 3 µm, a "sweet spot" is found. Within this range, the translational time scale is long, the rotational time scale remains short, and the particle remains in frame for a significant portion of the video capture. In this optimal zone, we have a robust region of time averaging periods ranging from 3 - 6 seconds that allow for ideal separation between the translational and rotational fluctuations. To account for smaller and larger particles, we selected a shorter averaging period within the working range of 3.5 seconds as our optimum setting allowing for flexibility in our downstream implementation.

#### 2.3.2 Non-linear rotational mapping for the recovery of θ(t)

From the prior section, we are able to extract the normalized intensity contributed by the IJP’s rotation *I_θ_*(*t*) by deconvolving the translational and rotational dynamics. Via windowed averaging and intensity normalization over all pairs of frames for a given time series, we are able to convert further (*I_θ_*(*t*), Δ*I_θ_*(*t*)) to Δ*θ*(*t*). Using weighted binning, we develop Gaussian distributions of Δ*θ*(*t*) for each particle track. The variance of these distributions provides a mathematical link to the rotational diffusivity *D_r_* and effective hydrodynamic diameter *d_p_* of a given IJP track per Eq. (2).

Both the isolated translational *I_Z_*(*t*) and rotational *I_θ_*(*t*) contributions towards the brightness time series are used to evaluate the approximate size of the particle *d_p_*. From Eq. (15), we approximated the particle’s translational distance |*Z*|(*t*) from the focal plane based on the *I_Z_*(*t*). This established distance allows us to weight different points along the time series by confidence. When the particle is in or near the focal plane, it appears brighter overall. Under these circumstances, changes in brightness from rotation yield a broader dynamic range from the camera’s sensor, thus providing significantly greater resolution. Since the video capture *DoF* is variable for different video capture set-ups, it is not easily derived. However, using the inverse of the dimensionless distance 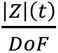, we develop a positional confidence heuristic that is independent of the explicit fitted parameter *DoF* and allows for particle-to-particle and video-to-video comparability.

Using this 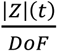 parameter, we weight brightness changes at each time along a particle path to preferentially value moments of high confidence over those of low confidence in the summary mean. Using the model of a theoretical particle (Figure 6a-b) alongside a similar workflow to that which was presented in Section 2.1, we simulate video capture of individual translationally-static, blinking, hemispheric IJPs of any size impacted only by rotational diffusion. Janus particles of nine diameters ranging from 500 - 5000 nm (*n* = 24 per size bin) were simulated with variable levels of intensity of the non-fluorescent side *α* = *I_Gold_*⁄*I_Fluor_* = {0.35,0.45,0.55,0.75}. Empirically fitting for *α*, the rotational contribution is renormalized by setting the curve *I_Gold_* = 0 and *I_Fluor_* = 1 per Eq. (16). In doing so, the isolated *I_θ_*_,*Est*_(*t*) grants us the approximate fluorescent intensity profile for an equivalent static particle in which the fluorescent side is perfectly bright and the gold side is perfectly dark. Applying the workflow detailed in Figure 7 – hereon referred to as the Culsans Method, we use *I_θ_*_,*Est*_(*t*) as the basis for estimating the size *d_p_* of a given particle via its rotational fluctuations. We mathematically describe the characteristic relationship of the simplified system by inverting Eq. (3) to yield the following.

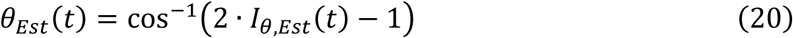

The above equation allows us to approximate the angular position *θ*(*t*) of the Janus boundary line between the bright and dim sides with respect to the image plane for a given video frame. To determine the angular difference between two frames, we look at the discrete-form of the derivative of Eq. (20), which diverges per 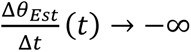 as *I_θ_*_,*Est*_(*t*) → 0 or *I_θ_*_,*Est*_(*t*) → 1. These conditions correspond to the “new moon” and “full moon” states, respectively. Under both conditions, Δ*θ_Est_*(*t*) steps for given Δ*I_θ_*_,*Est*_(*t*) steps become significantly non-linear and prone to estimation error due to non-linearity.

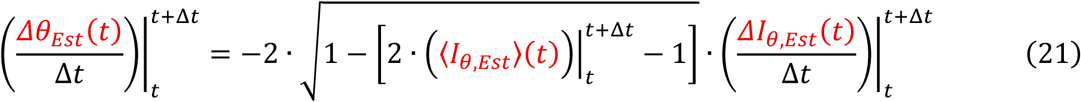

At a given time *t* for a time step of Δ*t* given by the framerate, 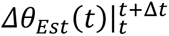 is evaluated using two unique terms derived from *ΔI_θ_*_,*Est*_(*t*) for every pair of frames. The first parameter is the instantaneous brightness 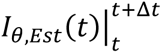 which is approximated as the average brightness 〈*I_θ_*_,*Est*_〉(*t*) of the particle between the two frames in consideration (Figure 7a-c). The second parameter is the fluorescent intensity change 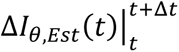 over the time step (Figure 7d-f).

**Figure 6.**
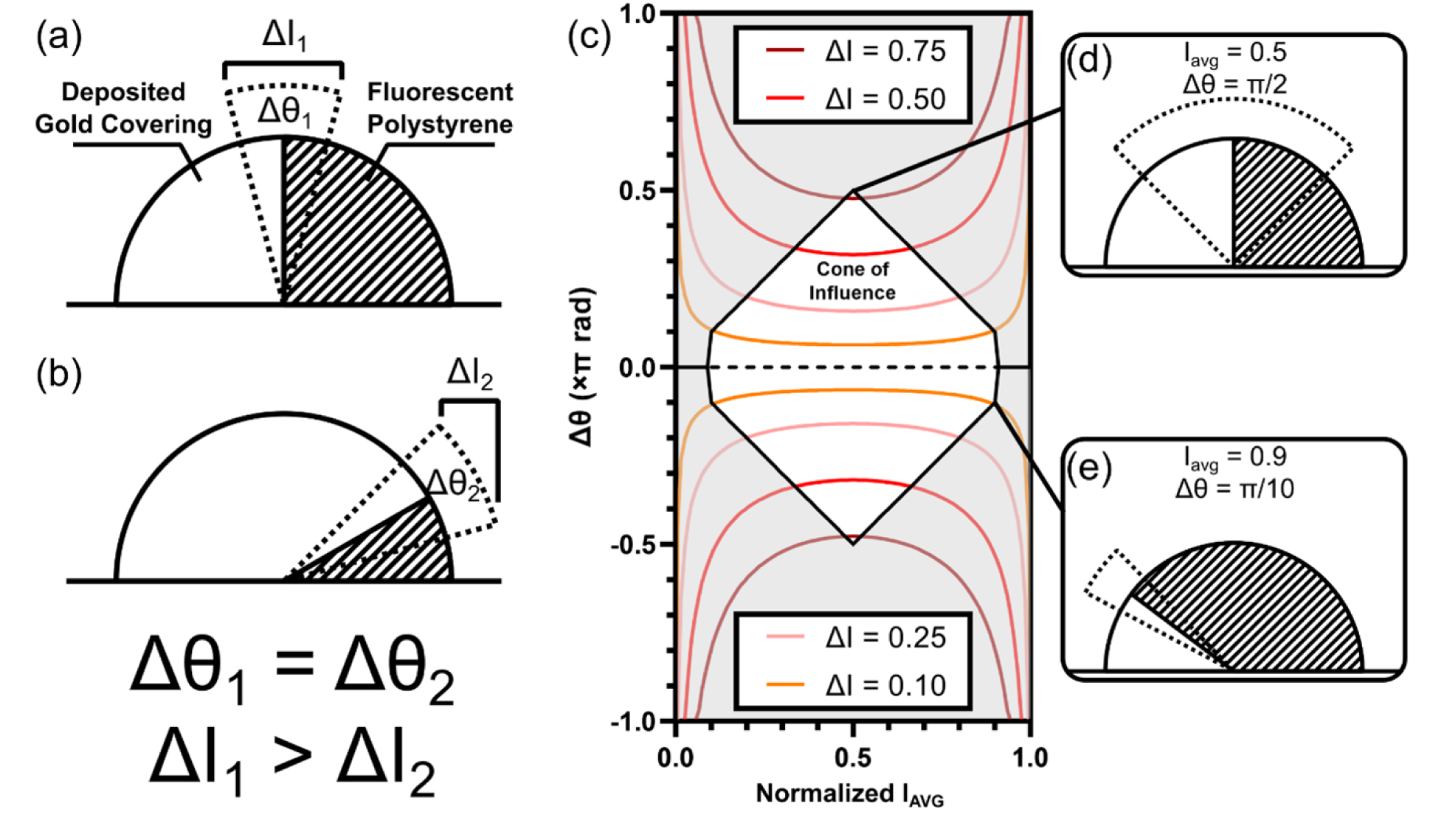
Non-linear conversion of normalized Δ*I* and *I_Avg_* to Δ*θ*. (a - b) Side-view representations of the observable side of two particles experiencing the same Δ*θ* deviation, yet (a) the conformation featuring the Janus boundary situated orthogonal to the image plane yields a greater Δ*θ* compared to (b) when the Janus boundary is off-skew. (c) Conversion phase map used to match measured normalized *I_Avg_* (horizontal axis) and lines of constant Δ*I*-values (red lines) to predicted Δ*θ* (vertical axis) values. Cone of influence (COI) assists in cleaning data by rejecting impossible values and low confidence values due to edge effects. (d - e) Side-view representations of conformers occurring at indicated regions in phase diagram.

**Figure 7.**
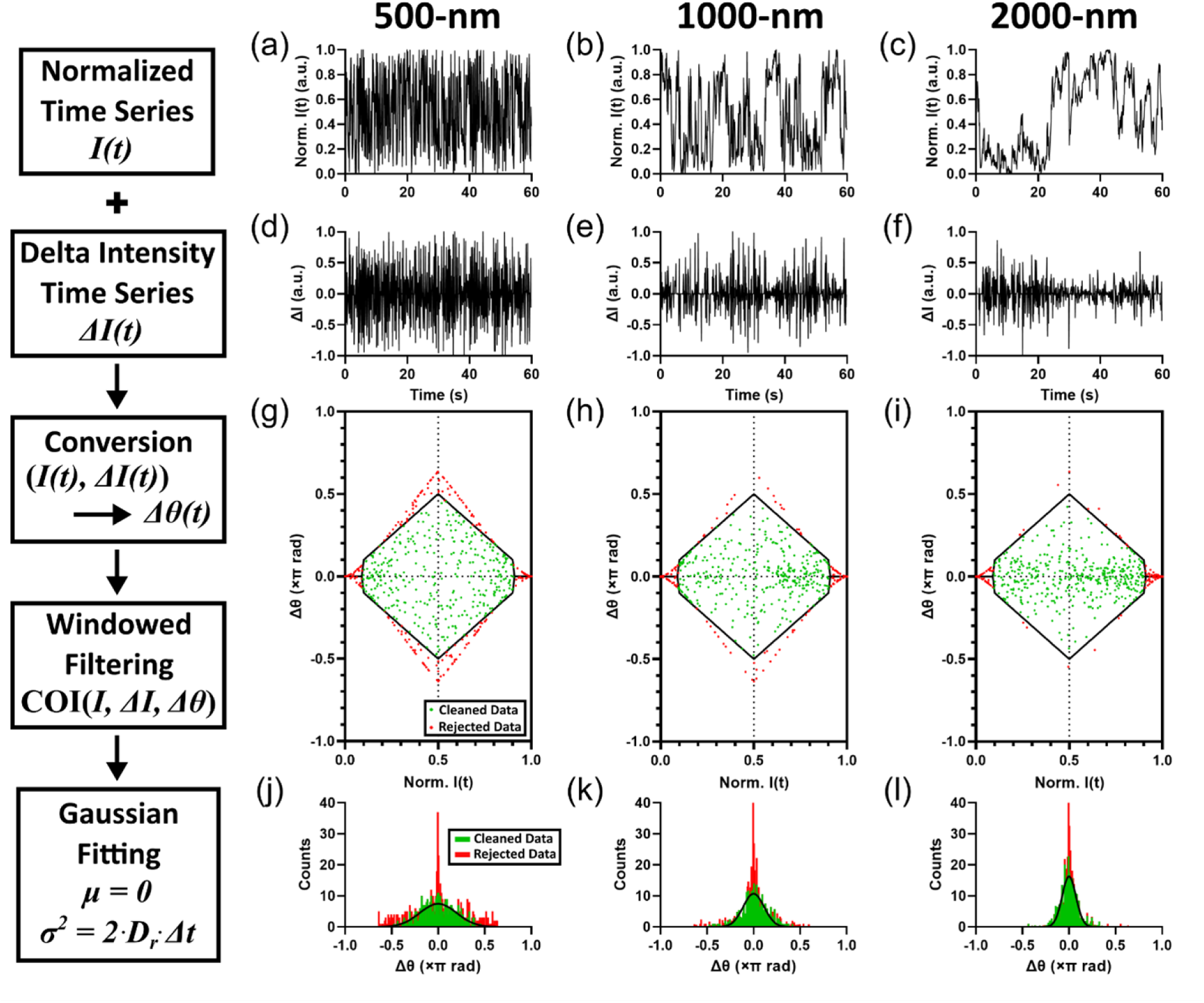
Representative signal processing workflows of the Culsans (Δ*θ*) Method for processing of tracked time series of three simulated static spherical IJPs of different sized diameters: (a, d, g, j) 500 nm, (b, e, h, k) 1000 nm, and (c, f, i, l) 2000 nm. Raw time series brightness data *I*(*t*) over the course of 1 minute (*t* ∈ [0,60] seconds) is (a - c) normalized to set the *I_Gold_* → 0 and *I_Fluor_* → 1 corresponding to minimum and maximum brightness values. (d - f) Individual brightness changes Δ*I* between adjacent frames at each time step (Δ*t* = 0.1 seconds) are calculated. (g-i) Using 〈*I_θ_*〉(*t*) and Δ*I_θ_*(*t*), (〈*I_θ_*〉, Δ*I_θ_*) → *θ* conversion phase maps are generated and cleaned per Figure 2c. Green data points represent retained, cleaned data, while red data points represent rejected data. (j - l) Cleaned data is binned by frequency counts for each given Δ*θ*(*t*) step. Green bars depict histogram of cleaned datapoints. Red bars are stacked counts of rejected data points outside of filtering window. Black line represents best fit Gaussian distributions for cleaned datasets.

The non-linear relationship is the result of the observed brightness being function of the projection of a 3D dihemispheric particle onto a 2D image plane rather than the option of the 3D particle itself. In this relationship between *I_θ_*(*t*), Δ*I_θ_*(*t*), and Δ*θ*(*t*), identical Δ*θ*(*t*) values yield drastically different scalar values of Δ*I_θ_*(*t*) based on different orientations of the Janus boundary with respect to the exposed particle hemisphere (Figure 6a-b). Plotting contours of constant Δ*I_θ_*(*t*) in the (〈*I_θ_*〉(*t*), Δ*θ*(*t*)) plane (Figure 6c), we generate a conversion phase map between these components. We find that the relationship between Δ*I_θ_*(*t*) and Δ*θ*(*t*) exhibits fairly stable linearity across ranges of 〈*I_θ_*〉(*t*) so long as the Janus boundary at the *t* and *t* + Δ*t* do not approach the 0 or *π* radians.

In the cases where nearly the entirety of the fluorescent side (〈*I_θ_*〉 → 1) or the entirety of the gold side is facing the observer (〈*I_θ_*〉 → 0), the non-linearity dominates and leads to difficulties differentiating the magnitude of a given Δ*θ* change. We define a Cone of Influence (*COI*) to systematically filter out Δ*θ*-values of known low confidence. When the Janus boundary is directly orthogonal (〈*I_θ_*〉 = 0.5), we limit Δ*θ* to be no greater than *π*⁄2 radians (Figure 6d). When the Janus boundary is close to the plane (*I_θ_*(*t*) ∈ [0.0,0.1] ∪ [0.9,1.0]), we reject everything within these regions as reported errors are on the scale of any potential signal difference (Figure 6e).

#### 2.3.3 Statistical fitting for hydrodynamic particle diameter d_p_

With the established non-linear correlation between (*I_θ_*, Δ*I_θ_*) and Δ*θ*, identical Δ*θ* values yield drastically different scalar values of Δ*I_θ_* (Figure 7a-c) based on different orientations of the Janus boundary with respect to the exposed particle hemisphere *I_θ_* (Figure 7d-f). We plot coordinate pairs of (*I_θ_*, Δ*I_θ_*) in the conversion phase map developed previously (Figure 7g-i). By binning evaluated Δ*θ*-values from all pairs of frames from a particle track that lay within the *COI*, we generate a histogram for all angular changes for a given rotational degree of freedom that occurs along the tracked trajectory (Figure 7g-i). Fitting a Gaussian distribution centered at *μ*_Δ*θ*_ = 0 (Figure 7j-l), we are able to estimate the diameter of the tracked particle *d_p_* via the quantifiable spread 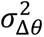 of the fluorescent flickering behavior over the particle track.

Per Eq. (2), the rotational diffusivity *D_r_* is related to the variance of the step deviations 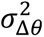 in a given singular degree of rotational freedom Δ*θ* by the given relationship 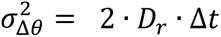 where 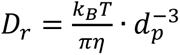. Intuitively, the correlation of 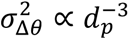 means that larger particles exhibit fewer extreme angular events along their trajectory paths for the same set of discrete time steps. We define large angular jumps as being events that exceed a sweep of ± *π*⁄4 radians, while small angular jumps are those that approach 0 radians and bounded by [− *π*⁄4, + *π*⁄4] radians. In our simulated particles, we observe that smaller particles experience more frequent large angular jumps in observed brightness level than do larger particles. In larger particles, angular jumps are rarer but not absent, leading to the observed brightness levels minimally fluctuating about a local equilibrium angle until a large enough thermal kick arrives to force the rotation to a new equilibrium angle. This behavior is reflected in the conversion phase maps and histograms where the larger particles have Δ*θ*-values that cluster tighter to 0 radians than those of the smaller particles.

With tighter histograms, the 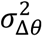-value becomes smaller which correlates to smaller *D_r_* – and, consequently, larger *d_p_*. The stacked histograms for each size exhibit steep, non-Gaussian peaks situated around Δ*θ* = 0 that are filtered out by the *COI* window. The pairs of frames tied to these points are associated with moments when either the IJP in the first or second frame in a pair approximates the “new moon” or “full moon” states as discussed earlier. In these instances, reliability of the correlation between 〈*I_θ_*〉, Δ*I_θ_*, and Δ*θ* begins to break down due to the non-linearity in the relationship. Additionally, edge effects attributed to the swing of the Δ*I_θ_* enters the fringe region described above are eliminated by the upper and lower bounds of the *COI* window. This *COI* filtration is crucial for the particle diffusivity *D_r_* approximation as we see that without this filtering step the Gaussian fitting completely fails to recover the expected particle diffusivity *D_r_* trend (Figure 8a). Additionally, the full workflow detailed in Section 2.2 applied to particles simulated with both rotational and translational transience maintains linearity for the same range of particle sizes (Figure 8b), which demonstrates the methodological robustness against non-ideal and transiently inconsistent data.

**Figure 8.**
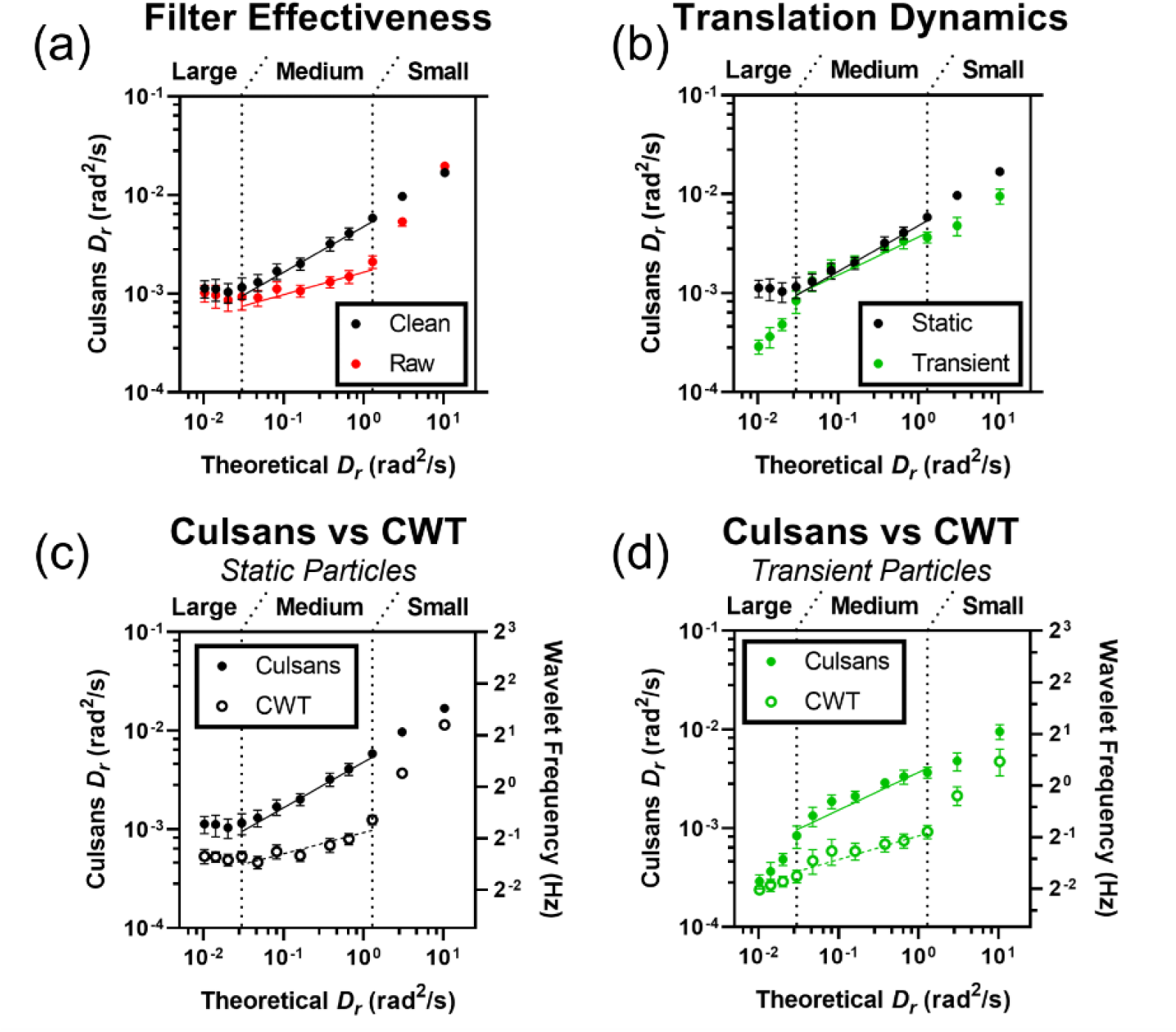
Comparative calibration curve sets demonstrating capability of the Culsans *D_r_* estimator to predict simulated particle diffusivities for simulated IJPs ranging in size from 500 to 5000 nm. Culsans *D_r_* estimators and CWT frequencies are plotted against theoretical rotational diffusivity *D_r_* values. Regions of Theoretical *D_r_* values corresponding to Small (500 - 1000 nm), Medium (1250 - 3000 nm), and Large (3500 – 5000 nm) particles are indicated in each figure. (a) Calibration series demonstrating effectiveness of COI filtering window showing the linearized predicted Culsans *D_r_* estimator values (black solid) against the unfiltered estimator values (red solid) for the translationally static simulated IJP data set. (b) Calibration series of filtered Culsans *D_r_* estimators for simulated translationally static (black solid) and transient (green solid) IJPs. (c - d) Comparative calibration series for the Culsans *D_r_* estimator (solid series) and the continuous wavelet transform (CWT) used in our prior work (open series) on simulated translationally (c) static (black series) and (d) transient (green series) IJPs. All error bars represent 95% confidence intervals. Each data point has *n* = 24 simulation replicates.

### 2.4 Comparison of predictive capabilities against Continuous Wavelet Transform (CWT)

The novel Culsans methodology exceeds the capabilities of the previous Continuous Wavelet Transform (*CWT*) approach.^12^ Using the Wavelet Toolbox from MATLAB, this technique allows for efficient detection of localized periodic behavior by fitting a “mother wavelet” with manipulable amplitude and wavelength. By scanning along these parameters, we generate magnitude scalograms describing periodic behavior both in frequency and time space. In our prior workflow, we used the Analytical Morlet wavelet to fit across the entire video duration using a range of dilated wavelet periods spanning 0.25 - 4.5 seconds. By taking the periods of best fit for each frame along the video and weighting them by their relative Fit Confidence magnitude, a weighted consensus period is calculated for each particle track as the key comparative metric for analysis. The full methodology (described in Supplementary Figure S2) provides several key strengths that account for many characteristics observed along particle tracks. Similar to the Culsans Method, local brightness weighting allows for emphasis of time points of high Fit Confidence. In contrast to the Culsans Method, the data processing is comparatively fast and efficient. The *CWT* approach completes analysis on a single particle track in 68 milliseconds, whereas the Culsans Method does so in 279 milliseconds.

Despite these benefits, the *CWT* approach inherently holds four crippling weaknesses. Firstly, the resultant weighted period metric lacks a clear theoretical connection to the claimed rotational diffusivity term *D_r_* beyond the tenuous claim of the units being that of inverse time (*s*^−1^). This definition omits the critical angular component in the full units (*rad*^2^⁄*s*) which sets the reference frame for comparison. Secondly, the *CWT* attributes all fluctuation behavior to rotational diffusivity despite clear *Z*-axial translational behavior. This attribution only works for smaller particles (< 1 µm). Small particles experience significantly reduced translational motion due to the 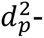 and 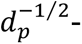scalings in ballistic gravitational settling and *Z*-axial translational diffusivity, respectively, as compared to the 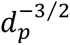 scaling in rotational diffusivity. Thirdly, the CWT method preferentially masks low period (high frequency) events by the dominant high period (low frequency) trends. As can be seen in Supplementary Figure S2e, the high period events are primarily contributed by *Z*-axial translation which will inherently exhibit a greater overall magnitude of change as the particle traverses through the entire focal volume.

Finally, the CWT always assumes to find periodicity in a given time series, which simplifies the interpretation of *D_r_* to that of a frequency term that scales with particle size by 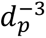. This assumption misinterprets the IJPs as spinning periodically like a planet when they are in reality jiggling in their angular orientation as a response to thermally applied torques from every direction. Once more, this misinterpretation holds for small IJPs (< 1 µm) as they experience more frequent rotational kicks between frames that span significant portions of their entire breadth of rotation 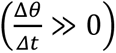 allowing a relatively faithful approximation of periodic rotation. This assumption fails for larger IJPs (> 1 µm) as they tend to primarily experience rotational kicks between frames of magnitudes significantly less than the available breadth of rotation 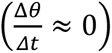. As a result, large IJPs display a relatively fixed orientation for extended periods of time. On the rare occasion that a large enough torque impulse spontaneously occurs, the *θ*-orientation of a large IJP may be shifted to another equilibrium for another statistically random amount of time. As the intervals between “jump” events are not regular, the behavior of the IJP becomes distinctly non-periodic. All together, we grasp that the CWT approach allows resolution of rotational behavior in smaller particles (< 1 µm) where translation does not significantly dominate, and where rotational fluctuations can be approximated as spinning. For larger IJPs (> 1 µm), the dominance of translational behavior completely eliminates any chance for isolating the rotational signal. In contrast, the Culsans Method is designed specifically to take advantage of translational forcing and fluctuation. This leads to a far more robust methodology that provides significantly improved linearity over a far broader range of observed behaviors, including those of IJP aggregates that will be further analyzed in the upcoming section (Figure 8).

We compare the calibration curves generated from the Culsans Method (solid series) and CWT Method (open series) run on rotating particles simulated as translationally static (Figure 8c) and transient (Figure 8d). As we discussed earlier, the CWT approach loses linearity for medium to large particles (> 1 µm) for both translationally static and dynamic IJPs. Meanwhile, despite the Culsans Method gaining in noise when processing translationally dynamic IJPs – as opposed to static IJPs, it maintains significant linearity within the entire dynamic range. This is reinforced by the Coefficients of Determination *R*^2^ for log_10_ − log_10_ fittings (Supplementary Table S1). These calibration curves demonstrate that the well-defined *COI* filtration process directly facilitates the predictive capabilities of the Culsans Method. Further, the Culsans Method provides a significantly more linear calibration curve (*R*^2^ ≥ 0.85) than the CWT Method (*R*^2^ ≤ 0.64), meaning that the new analytical method yields a statistical model that explains significantly more of the variation observed in the data. As such, the linear calibration fitting for the translationally dynamic dataset using the Culsans method is used to interpret empirically observed data for the remainder of the study.

### 2.5 Quantification of low-concentration virus by leveraging IJP agglutination

To further extend the applicable domain of the IJP platform, we test the efficacy of the new analytical workflow with the detection of low concentration viruses in realistic biofluid. IJPs were functionalized with anti-SARS-CoV-2 Spike Glycoprotein antibodies and incubated in human plasma spiked with serial dilutions of PCR-validated genome equivalent (GE) concentrations of heat-inactivated SARS-CoV-2 virions (Figure 9). The anti-SARS-CoV-2 IJP platform was tested against concentrations of target previously unbreachable using the CWT approach (Figure 10e-f). With the improved Culsans analysis, significant improvement (Figure 10a-d) is achieved over our established Anion Exchange Membrane (AEM) biosensor technology^14–17^ with LOD of 10^6^ - 10^7^ particles/mL (Figure 11). For more information on the AEM biosensor platform developed for this application, see Supplementary Figure S4. Enhanced by the new Culsans analytical methodology, we found the sensitivity of the IJP assay for SARS-CoV-2 to be highly sensitive to the ratio of viral particle concentrations to IJP concentrations *λ*. For the Poisson limit – also known as the “Rare Event Limit” where *λ* < 1, the distribution of effective hydrodynamic radii *d_p_* for a given population of IJPs approaches a narrow band around an expected diameter of 1 micron (Figure 10c). However, when *λ* > 1 known as the “Gaussian Approximation Limit”, every IJP within the sample will be bound to a number of viral particles distributed in accordance to a Gaussian normal model.

**Figure 9.**
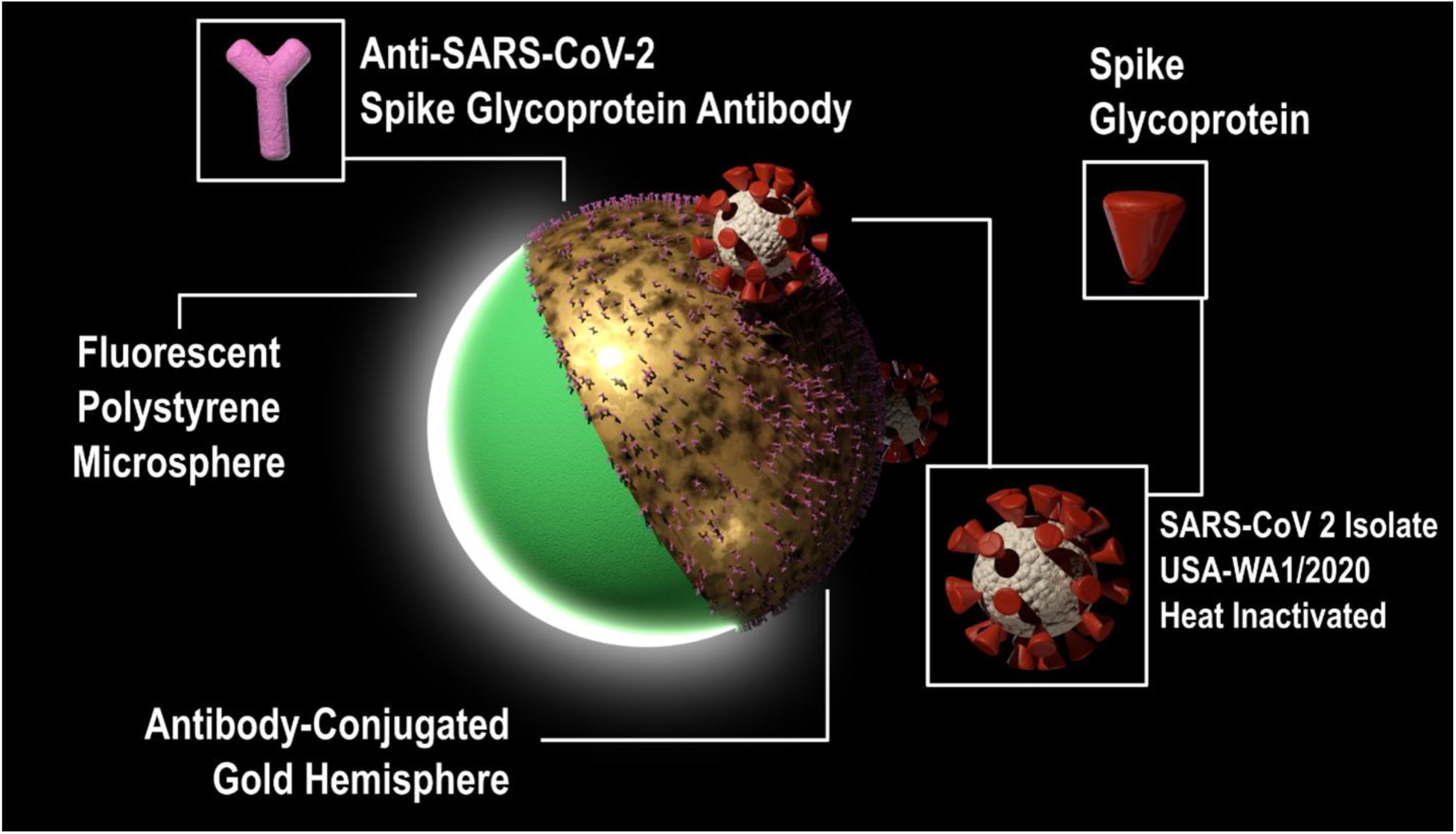
Depiction of Anti-SARS-CoV-2 IJP platform developed in this work to quantify low concentration viral content in human plasma. Fluorescent IJPs were functionalized using Anti-SARS-CoV-2 Spike Glycoprotein antibodies. When these particles are spiked into filtered plasma containing low-concentrations of heat-inactivated SARS-CoV-2 viral content, the IJPs bind to the available viral target which cause their characteristic Brownian rotation behavior to change.

**Figure 10.**
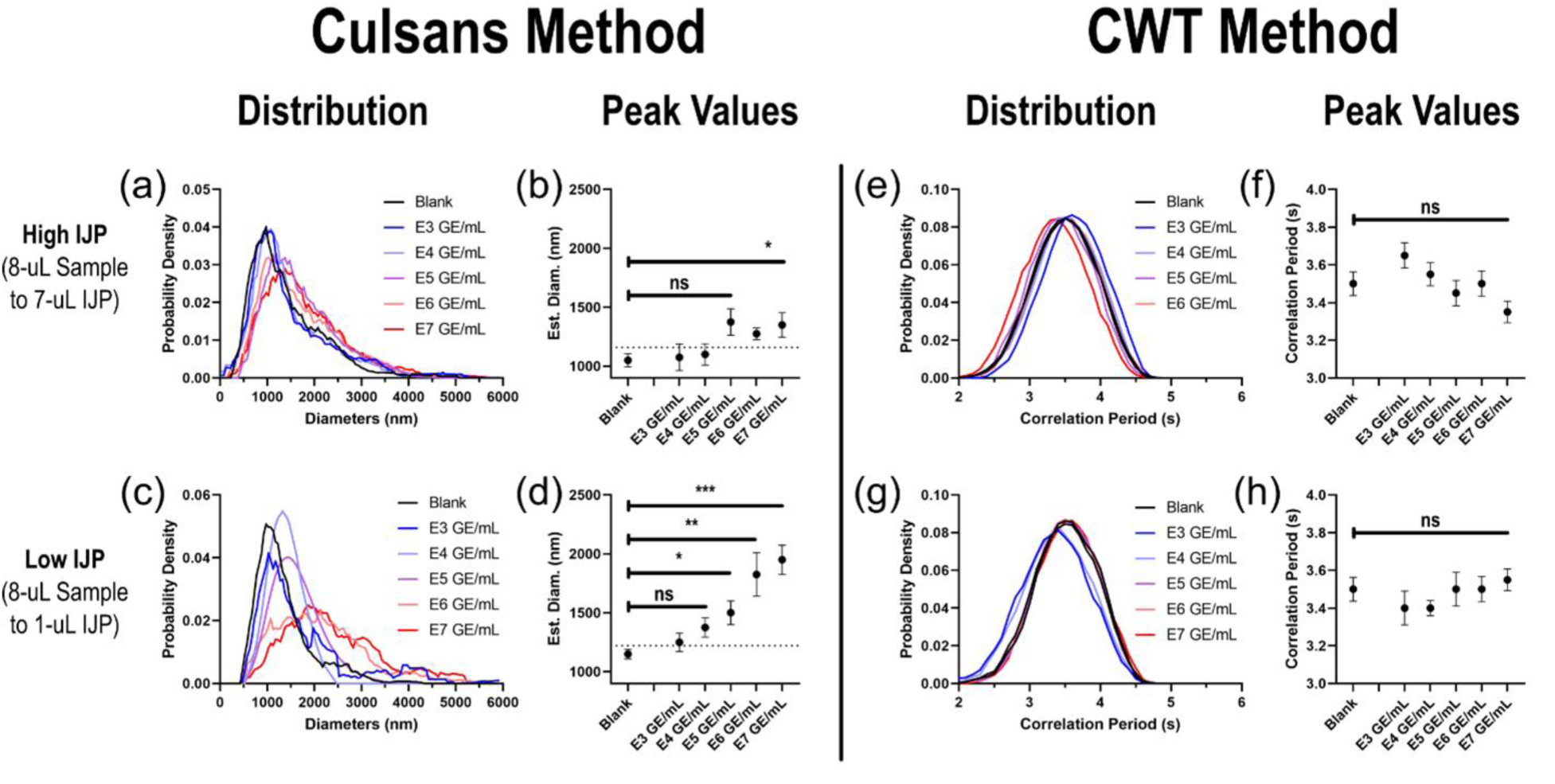
Comparative distributions and peak value calibration curves of IJP biosensor readouts analyzed using (a - d) the novel Culsans Method and (e - h) the CWT Method. Top row corresponds to experiments using standard IJP experimental composition of ∼10^6^ particles/mL with varying genome equivalent (GE) virus concentrations. Bottom row corresponds to experiments using reduced IJP conditions (∼10^5^ particles/mL) significantly increasing the Virus-to-IJP ratio. Note the long tails in the IJP aggregate size distribution detectable only by the Culsans method but not the CWT method. Calibration curves were generated using the smoothed maxima (peak) of each distribution with error bars representing the region about the peak corresponding to value greater than 90% of the maximal probability.

**Figure 11.**
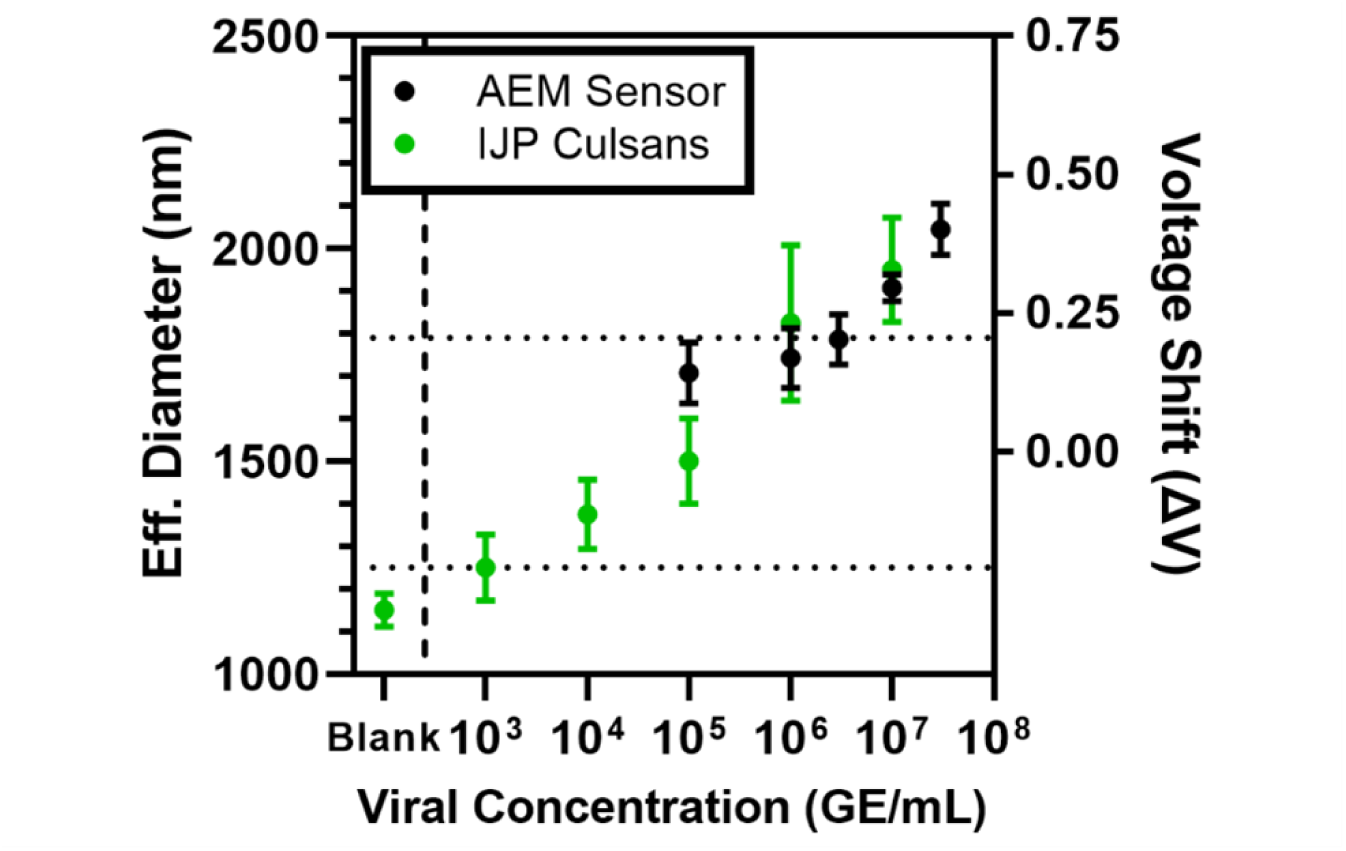
Overlay of Anti-SARS-CoV-2 glycoprotein calibration curves generated using IJP platform (green series) and Anion-Exchange Membrane (AEM) biosensor platform (black series) for orthogonal validation and benchmarking. The limit of detection (LOD) for the AEM is calculated to be about 10^7^ GE/mL of genome equivalent (GE) viral content, whereas the *LOD* for the Culsans processed IJP platform is calculated to be about 10^3^ - 10^4^ GE/mL of viral content. Concentrations below the LoDs could not be discerned using each respective platform.

Most interestingly, under low virus to IJP conditions (*λ* ≤ 1), nearly all the IJPs are undocked and empty. The size distribution would be dominated by the signals contributed by unbound IJPs, and any signal from the rare IJP-virus conjugates would be drowned out. This would typically define a lower bound *LOD* for most biosensors. However, the IJP platform allows us to adjust the IJP concentration in accordance to the relative target virus concentration to further improve the *LOD*. This tunable parameter (*λ* = *Virus*: *IJP*) significantly enhances the population of IJP-virus conjugates with respect to unbound IJPs.

Recognizing this relationship, we design experiments with two specific IJP concentrations and adjust the virus concentrations to get conditions with matching *λ* at different virus concentrations. Typical experiments have been performed at 10^6^ IJP/mL (hereon referred to as “High IJP”), while reduced IJP experiments were run at 10 × lower IJP concentration (referred to as “Low IJP”). When the experiments are run under “Low IJP” conditions, we gain significant improvements in detection sensitivity. The calibration curve yielded with the Culsans analytical method gains significant linearity down to 10^3^ - 10^4^ GE/mL (Figure 10c-d), where GE denotes genome equivalent viral count. We clearly see the change in particle distribution with increasing concentrations of target viruses. Even with the CWT method, we gain linearity and a minor positive (though non-significant) correlation between target concentration and wavelet correlation period (Figure 10g-h).

For increasing virion concentrations, we see a distinct shift in the detected particle size distribution. The estimated diameter trends to almost 2 microns, nearly double to original particle size. The scale of the size shift is much larger than what can be attainable with virions docking to the surface of IJPs alone. As IJP concentrations are held constant in both experimental schemes, we may better interpret this behavior by normalizing our calibration curves for both experimental conditions against each incubation IJP concentration. Doing so, both series may be compared against a common independent variable and derive a calibration with respect to the Poisson *λ* (Figure 12a). For both the "High IJP" and "Low IJP" experiments, calibration curves converge with respect to a common *λ* variable where significant signal change begins upon reaching *λ* ≥ 10^−1^.

**Figure 12.**
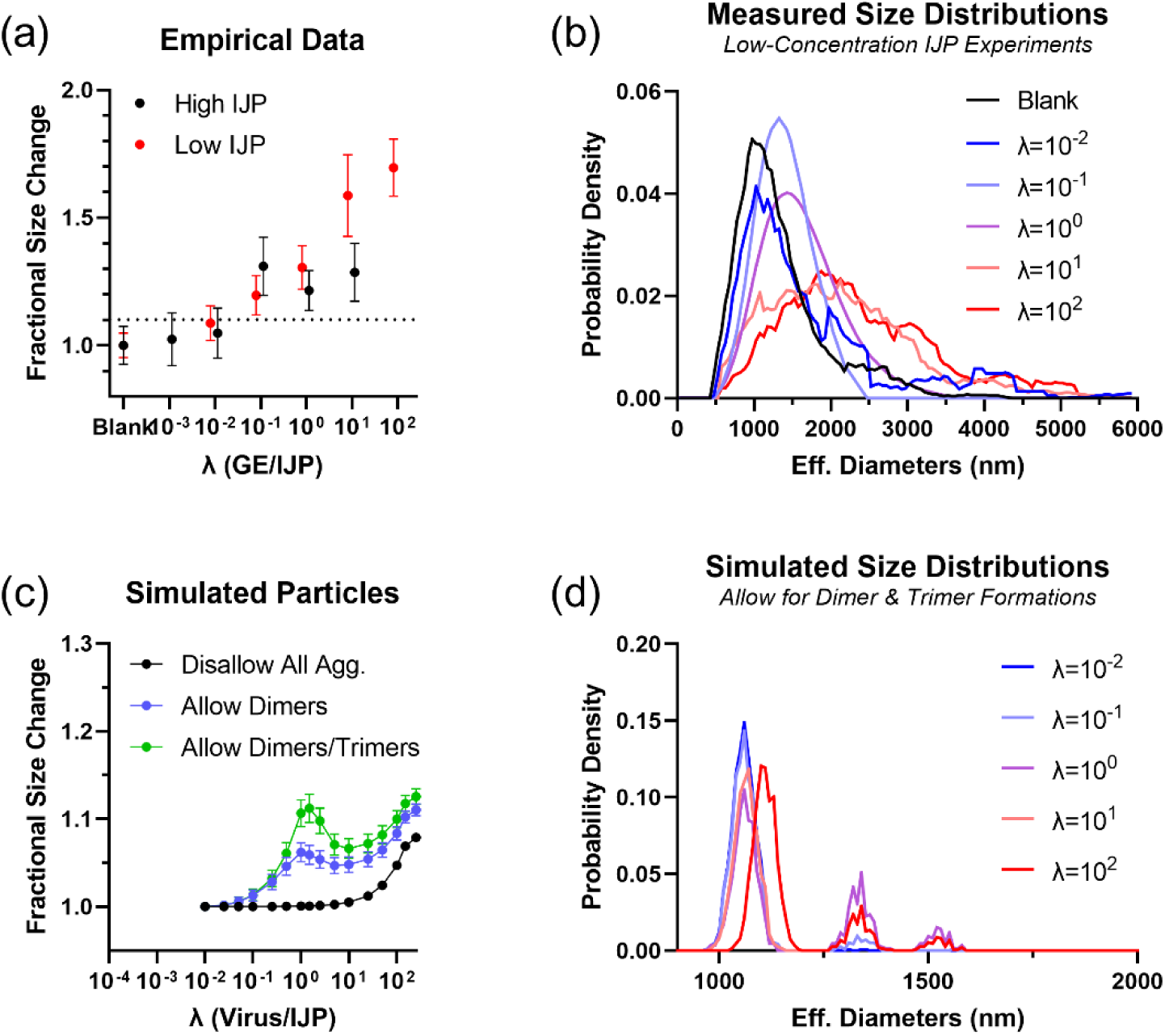
Leveraging Poisson regime amplification via targeted aggregation to facilitate low-concentration particles detection. (a) Fold change calibration curves derived using the Culsans Method normalized by IJP concentration for both the “High IJP” and “Low IJP” experimental conditions. Lambda (*λ*) represents the relative viral genome equivalent (GE) concentration in solution per IJP present. A local maximum in the fractional size change is observed at *λ* = 1, when the viral and IJP concentrations match. This is followed by a local minimum for increasing *λ* which yields a distinct non-monotonic inflection in the fractional size distribution (b) as demonstrated in the effective particle diameter distributions for the “Low IJP” experimental set. (c) Calibration plot and (d) size distributions for simulated particles (*n* = 120 per *λ* distribution). Lambda (*λ*) represents the relative viral particle concentration in solution per IJP present. The maximum in fractional size change at *λ* = 1 is only observed when agglutination events are allowed. (c) Three aggregation cases were simulated with the cases of (black series) allow no aggregation assuming only viral capture events, (blue series) allow IJP dimerization assuming only IJP-Virus-IJP sandwich agglutination possible, and (green) allow IJP dimerization and trimerization events. All binding and aggregation events are assumed to be irreversible due to multi-valent virus-IJP binding.

To interrogate the mechanistic feasibility of this detection limit, we used a numeric population simulation of IJPs (1 µm) binding to Poisson distributions of target nanoparticles (0.2 µm). For each capture event, the simulated particle would have its diameter increased by an effective measure based on the total volumetric difference between the original IJP and the new IJP-target conjugate.

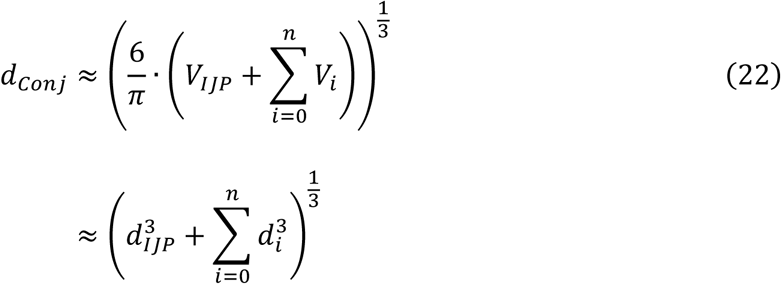

where *d_Conj_* is the approximated effective hydrodynamic diameter of the composite IJP-target conjugate and *V_i_* and *d_i_* are the individual volume and diameter, respectively, of each of the *n* targets attached to the same IJP for each binding event it experiences. For the classical case where the only behavior is virions binding to IJPs, no significant size changes occur until after *λ* ≥ 10^1^ (Figure 12c black series and Supplementary Figure S7a). This result suggests the existence of an additional mode of binding that can significantly and irreversibly increase the particle size.

We propose the mechanism of IJP aggregation due to multi-valent virus binding (known as agglutination) wherein the joining of two or more IJPs is achieved via the bridging of shared viral targets. A focused algorithm was developed to simulate dimerization events by pairing and joining IJPs that experience exactly one binding event via Eq. (22) (Figure 12c blue series and Supplementary Figure S7b). (To simplify calculations, aggregates were simulated as single IJPs of comparably increased size.) Likewise, trimers were generated for groups of IJPs that each individually experience exactly two binding events (Figure 12c green series and Supplementary Figure S7c). These agglutination events are likely as, when in the Poisson limit, an existing virion-IJP conjugate is more likely to encounter a blank IJP than another unbound virus. Alternatively, IJPs that experience ≥ 3 binding events are most likely to be within the Gaussian Approximation limit. Under these conditions, we assume that each virion-IJP conjugate is more likely to encounter many more unbound viruses than bare IJPs. In fact, as the total number of sites per IJP is filled up, it is less likely for a conjugate to subsequently dimerize with another conjugate. The corona of bound viruses on each IJP will further sterically hinder and prevent aggregation events leading to convergence towards the classical regime of virion-IJP singlets dominating.

As noted earlier, the simulated population mean only shifts significantly for *λ* ≥ 10, in the case of no bridging events. Yet, for the cases where aggregation is allowed, the population mean begins shifting for much smaller *λ* > 0.1. This earlier shift in the population mean helps provide an avenue to explain our significantly enhanced LOD using the IJP Culsans methodology. Rather than just detecting virion-burdened IJP size changes, we are able to also pick up virion-bridged IJP-IJP aggregates within the Poisson limit.

To definitively pin down that this aggregation behavior, we incubated streptavidin conjugated IJPs with concentrations of 200 nm biotinylated polystyrene (PS) nanobeads spanning a range of *λ* ∈ [10^−2^, 10^2^]. Using Field-Emission Scanning Electron Microscopy (FESEM), the samples were imaged in broad-field (at 500 × magnification) and near-field (at 2000 × magnification). The choice to use Biotin-SA chemistry and PS targets is to ensure bond stability throughout air-drying procedure and under vacuum conditions of SEM imaging. Air-drying has been reported to have significantly adverse effects on antibody stability due to conformational degradation of the delicate protein^18^, while Biotin-SA bonds remain stable throughout the process^19^. These effects are differentiated even further under vacuum conditions where antibodies completely degrade^20^ while Biotin-SA bonds become even stronger^21^.

Aggregates of varying numbers of IJPs were manual binned for near-field images (*n* = 10 per condition), while automated particle detection, area normalization, and size binning were performed for the broad-field images (*n* = 3 per condition) (Supplementary Figure S6). Clusters containing seven or more IJPs were subsequently rejected as those are likely to be due to off-target effects or contaminant binding. The fraction of clusters that represent IJP Singlets are compared to those that are aggregates of two or more IJPs in Figure 13. (For more detailed breakdown, see Supplementary Figure S5 and S9.)

**Figure 13.**
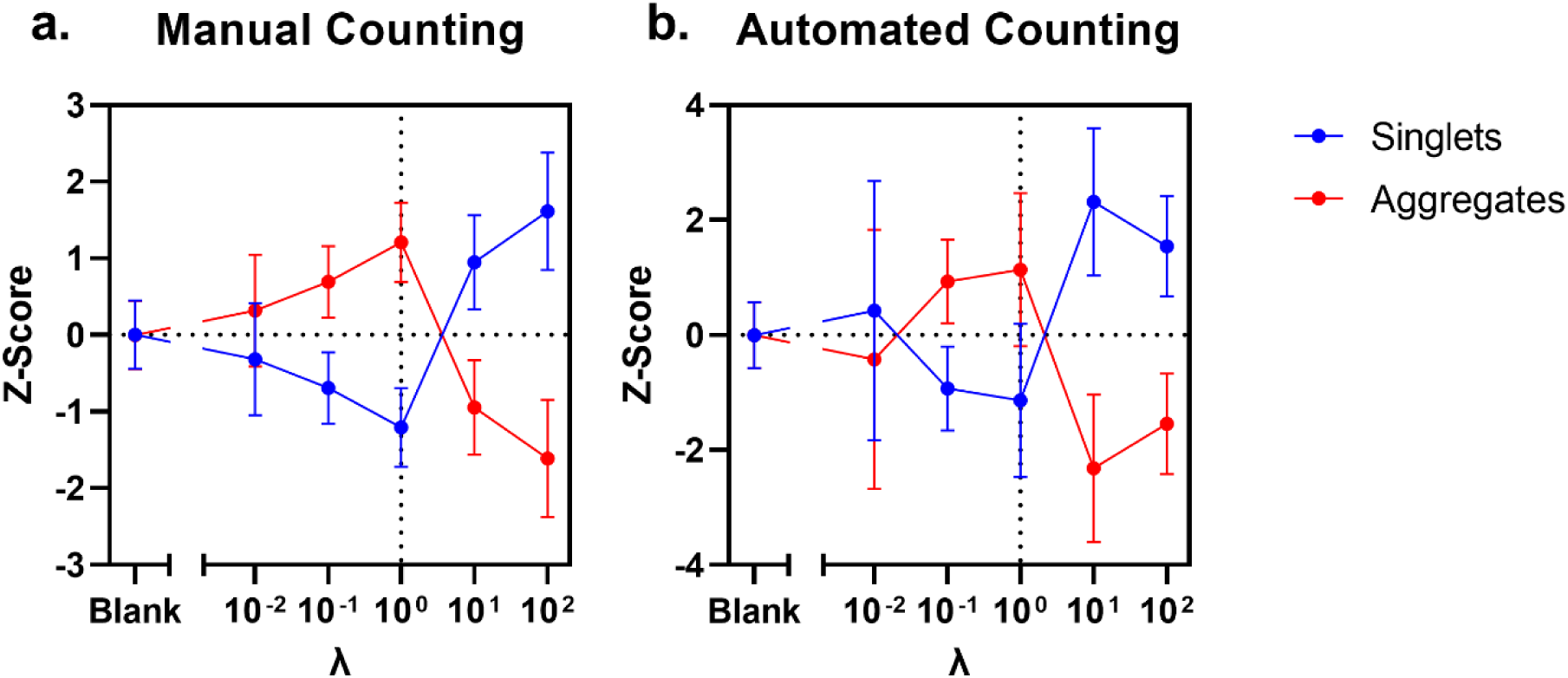
Relative prevalence of IJP singlets and aggregates for varying ratios of 200 nm biotinylated nanobeads to 1 μm streptavidin-functionalized IJPs incubated together, dropcast onto glass slides, and air dried. Prevalence values are expressed as Z-Score deviations from the mean of the IJP sample incubated without nanobead presence. Aggregates included clusters representing two to six total IJPs. Larger clusters are attributed to off-target contact-line aggregation effects. (a) Population results of near-field SEM images derived using manual aggregate counting and binning. Each data point represents singlet and aggregate counts from ten independent images. (b) Population results of broad-field SEM images derived using automated particle detection. Each data point represents singlet and aggregate counts from three independent images. Notice the reduction in singlets and gain in aggregates with increasing *λ* → 1, and subsequent increase in relative IJP singlets and decrease in IJP aggregates as *λ* ≫ 1. Agglutination is most pronounced at *λ* = 1. Error bars represent the standard errors of the mean between images.

A distinct bifurcation of behavior occurs after *λ* > 1, as shown clearly within Figure 13. When *λ* ≤ 1, the fraction of counted clusters that are IJP singlets increase in favor of more IJP aggregates. Moreover, after the inflection point when *λ* > 1, the population of IJP singlets reestablishes and becomes dominant again pointing towards the steric saturation discussed for the Gaussian Approximation limit. We propose that these striking behaviors are indicative of the aggregation mechanism we detailed in our simulations.

Interestingly, the results show that even in the Blank case significant quantities of aggregates exist. Yet, each set of IJPs is always thoroughly dispersed via sonication prior to incubation meaning it is unlikely for such an extent of aggregation to occur across the board under all conditions. This extensive aggregation is likely formed due to contact line particle chaining during the air-drying process^22^. (Supplementary Figure S8 provides examples of chained aggregates in IJP Sample without nanobeads.) These chains are unlikely to form through the bridging mechanisms as only a single hemisphere on each particle is capable of target-specific binding activity. Therefore, these chained aggregates exhibit length and directionality suggesting that the likeliest mechanism is this forced aggregation by capillary action at the liquid contact line.

Consequently, this means that all particle distributions will always skew towards aggregates over singlets. However, this skew is deterministic in that – if one population already consists of more aggregates than another population – the population with more initial aggregates will skew even further towards larger aggregates than the other population. Since larger aggregates consume more of the singlet fraction, we still see significant shifts in singlet-aggregate population differences in the resultant dropcast distribution. Taken as a whole, despite the distribution not being quantitatively representative of the expected population size changes seen in Figure 12, the underlying patterns and the distinct inflection point where aggregation dynamics shift (*λ* > 1) remain consistent and qualitatively solidify our confidence in our described mechanism.

Given this exploration, we may explain the mismatches in behavior of overall intensity of the shift in predicted size distribution despite the overlap in critical *λ* observed in Figure 12a. At the onset of the distribution shift (*λ* = 0.1), the size shift if “High IJP” trials is higher than that in the “Low IJP” trials. As the higher IJP concentration means that the likelihood of IJP-IJP dimers is increased, the subsequent signal shift is also increased. For increased viral concentrations beyond the inflection (*λ* > 1), the lower particle size distribution for the “High IJP” trials compared to the “Low IJP” trials can be explained as with more available IJPs in solution the surface saturation effect by locally available virions displacing IJP-IJP pairings becomes more rapid and uniform. This sort of “corona-forming” behavior (known as the “Hook Effect”) is well described in their ability to stabilize colloids and preventing aggregation^23^. This explains the drop in particle size in the “High IJP” case. For the “Low IJP” case, the same *λ* regime exists at a lower overall concentration of IJPs and virions, leading to a slower and less uniform saturation behavior. This bulk dilution approach to counteract and improve has also been historically discussed in the literature lending further support for our approach towards dynamic range improvements^24^.

## 3 Discussion

Here, we have adapted the IJP platform, previously utilized for cancer differentiation^12^ and staging^13^, towards a distinctly different clinical context consisting of a quantitative virus agglutination assay for low-count, early-stage, pretreatment-free, and in-field detection. Rather than focusing on single IJP rotation, we re-engineered the core computational framework to maximize information extraction from agglutinated aggregates. These agglutination events demonstrate significantly increased likelihood under low viral counts (by three orders of magnitude) than in high target concentration regimes. In recognizing the limitations of our previously applied CWT approaches that inaccurately assume signal periodicity, we recontextualized the system using deterministic and stochastic physics to exploit the rotational diffusivity inherent to these micron-scale particles and their aggregates. This framework led to an algorithm that deconvolutes complex signal streams that fractionate background noise to isolate statistically random rotational dynamics from similarly random translational dynamics.

From this, we derived a non-linear mapping (Culsans Method) to directly extract rotation diffusivity from fluorescent signal that yields significantly improved linearity in comparison to the CWT approach within the range of relevant particle sizes. Using the framework, we challenged our analytical platform against realistically low viral count samples (as low as 10^3^ copies/mL) in untreated plasma, previously only achievable with lab-bound RT-PCR techniques. We subsequently deduced a framework for a shifting dynamic range by controlled reduction of binding sites to eliminate the noise of unbound IJPs. This advantage is unique to the IJP platform due to the ease at which IJP concentration can be adjusted through dilution and is facilitated by the sensitivity of the Culsans method in its ability to report individual IJP behavior. We further revealed an aggregation mechanism that explains the exceptional LOD achieved. This behavior would not have been reliably measured had we not: (1) implemented the Culsans analytical framework, and (2) discovered how to shift our sensor’s dynamic range to enhance agglutination. Put together, we were able to extend of LOD down to 10^3^ - 10^4^ particles/mL allowing us to detect > 70% of total cases^1^ and also have the sensitivity to detect early infections, while using a platform that is rapid, requires minimal equipment, and uses low volumes of sample.

In summary, the derivation of the stochastic physics of IJP rotational diffusion provides a definitive mechanism for achieving single-particle sensitivity and quantifying aggregation-enhanced signal amplification. Together, these findings demonstrate how this application of the IJP technology can fulfill both the requirements for target sensitivity and deployment tempo necessary during large-scale viral outbreaks.

## 4 Methods

### 4.1 Immuno-Janus Particle (IJP) virion plasma spike-in experiment: gold functionalization, sample preparation, and video capture

IJPs were fabricated and functionalized with Anti-SARS-CoV-2 spike glycoprotein antibody - Coronavirus (Abcam, Cat. No. ab272504) per the methodology detailed in our prior works.^12,13^ Samples of SARS-Related Coronavirus 2, Isolate USA-WA1/2020, Heat Inactivated (BEI Resources, Cat. No. NR-52286) were diluted in 1 × PBS to working concentrations. Functionalized IJPs were washed three times in 0.1 × PBS and resuspended in 1 × PBS. IJPs were sonicated for 30 seconds prior to use. For the standard experiments, 8 µL of diluted viral samples were incubated with 7 µL of IJP stock solution. For the reduced IJP experiments, 8 µL of diluted viral samples were incubated with 1 µL of IJP stock solution. Samples were incubated on an orbital shaker at 750 RPM for 45 minutes. Each sample was imaged and particles fluorescent time series were tracked following methodology detailed in our prior works.^12,13^

### 4.2 IJP-nanobead agglutination experiments: streptavidin functionalization, bead capture, and SEM imaging

Fabricated IJPs were functionalized with Streptavidin (SA), lyophilized (Thermo Fisher, Cat. No. 434301) via EDC/NHS conjugation to the polystyrene (PS) hemisphere. Fabricated IJPs were washed and transferred into 0.1 M MES buffer, through centrifugation at 15 kxg. The resulting pellet was resuspended in activation solution containing 100 mM EDC and 25 mM NHS in 0.1 M MES (pH 6.0) buffer using bath sonication. This mixture was incubated for 30 min while mixed on an orbital shaker. Excess activation reagents were removed through centrifugation and resuspension in 1 × PBS buffer. The activated IJPs were resuspended and incubated with 0.1 mg/mL SA in 1 × PBS buffer for 8 hours in 4°C under continuous mixing to produce SA-conjugated silica nanoparticle reporters. The functionalized IJPs were then quenched in 100 mM Tris-HCl (pH 7.4) and incubated for 30 minutes while mixed on an orbital shaker. Functionalized IJPs were then three times washed and transferred into 1 × PBS buffer through centrifugation at 15 kxg for storage in 4°C until use.

Mixtures of 200 nm biotinylated polystyrene nanobeads (Nanocs, Inc., Cat. No. PS200-BN-1) were produced for a range of dilutions in DI H_2_O. SA-functionalized IJPs were washed and resuspended in DI H_2_O. Nanobeads were incubated with IJPs in a 1:1 ratio by volume on an orbital shaker for 45 minutes. Mixtures were dropcast onto labeled glass slides that were cleaned with isopropyl alcohol, plasma-treated, and mounted onto SEM stubs. The slides were allowed to dry overnight in a still-air box.

This work was carried out in part in the Notre Dame Integrated Imaging Facility (NDIIF) Advanced Electron Microscopy Core, University of Notre Dame, using the Magellan Digital Field Emission Scanning Electron Microscope (FESEM). The dried dropcast samples were sputtered with 2 nm of Iridium using the 8004 Desktop High Resolution Coating System for FE-SEM from Ted Pella, Inc. from the Notre Dame Integrated Imaging Facility (NDIIF) core facility. Samples were imaged using the Magellan FESEM run at 0.10 nA and 10.00 kV. Aggregates in all images were evaluated using ImageJ with a binary erosion approach and the Analyze Particles function. Images were thresholded for particle borders. Each particle was filled and eroded by 7 pixels.

### 4.3 Anion-Exchange Membrane (AEM) orthogonal virion biosensing

We offer a brief description of the AEM biosensing method below. More in-depth analysis and validation can be found within our dedicated papers on AEM sensing of lipoproteins, extracellular vesicles, and nanoparticles.^15–17,25–28^

A 9-well biosensor array holder and buffer reservoir were fabricated using a stereolithography 3D printer (Elegoo Saturn 3). Circular anion-exchange membranes with 1 mm diameter (Mega a.s., Czech Republic) were embedded in fast-cure polyurethane casting resin (TAP Quik-Cast System, TAP Plastics) using a custom-fabricated silicone mold. Following curing, the casted AEM sensors were mechanically inserted into the array holder to establish a sealed configuration.

Each sensing well consisted of a rectangular sample loading window and two circular electrode ports corresponding to the reference (R) and counter (C) electrodes. The central well served as a common reference compartment containing the working (W) and working sense (WS) electrodes. A Gamry Reference 600 potentiostat (Gamry Instruments) was used to apply electrical current between the working and counter electrodes across the AEM while simultaneously monitoring the voltage drop between the working sense and reference electrodes.

The AEM surface was first functionalized through UV-assisted grafting of carboxylic acid groups. Briefly, membranes were exposed to 200 mM 3,3’,4,4’-benzophenonetetracarboxylic acid dissolved in 0.1 M NaOH under ultraviolet irradiation.

Following functionalization, membranes were thoroughly rinsed with DI H_2_O to remove residual reagents and subsequently incubated in 0.1 M MES buffer (pH 4.7) for 2 hours at room temperature.

For carbodiimide coupling, membranes were incubated in activation solution containing 100 mM EDC and 25 mM NHS in 0.1 M MES buffer for 30 minutes. The activation solution was removed, and the membranes were immediately incubated with 0.1 mg/mL Anti-SARS-CoV-2 spike glycoprotein antibody - Coronavirus (Abcam, Cat. No. ab272504) for 8 hours in 4°C to enable covalent immobilization.

Silica nanoparticle reporters (50 nm, carboxyl-functionalized) were functionalized using a similar EDC/NHS conjugation strategy. Silica nanoparticles were first transferred into 0.1 M MES buffer, through centrifugation at 15 kxg to remove the stock storage buffer. The resulting pellet was resuspended in activation solution containing 100 mM EDC and 25 mM NHS in 0.1 M MES buffer using bath sonication and incubated for 30 minutes under continuous orbital mixing at 250 RPM. Excess activation reagents were removed through centrifugation and resuspension in 1 × PBS buffer. Finally, the activated silica nanoparticles were resuspended and incubated with 0.1 mg/mL SARS antibody for 8 hours in 4°C under continuous mixing to produce antibody-conjugated silica nanoparticle reporters.

### 4.4 Simulations and statistical analysis

Low intensity simulations were run using local Python infrastructure with JupyterLab. Additionally, this research was supported by the Notre Dame’s Center for Research Computing (CRC) through the use of their High-Performance Computing (HPC) cyberinfrastructure allowing for extended, memory-intensive Python simulations. The data presented here is expressed as the mean and standard deviation or standard error of the mean as calculated using Excel, Python, or GraphPad Prism 8 software functions. Errors are propagated using standard propagation of uncertainties techniques.^29^ The simulation and data analysis scripts and spreadsheets of this study will be provided upon reasonable request.

## Data Availability

All data produced in the present study are available upon reasonable request to the authors.

## 5 Acknowledgements

This work has been supported by the National Institutes of Health (NIH) 1R21AI180713-01A1 and the Berthiaume Institute for Precision Health (BIPH) at the University of Notre Dame Bioscope RFP fund. We would like to acknowledge the Notre Dame Integrated Imaging Facility (NDIIF) at the University of Notre Dame. We acknowledge BEI Resources as the source of and the U.S. Centers for Disease Control and Prevention (CDC) as contributor of the SARS-Related Coronavirus 2, Isolate USA-WA1/2020, Heat Inactivated (BEI Resources, Cat. No. NR-52286) sample used in this study. We agree that ATCC may inform the U.S. CDC of our identity if required to do so by law, by the U.S. CDC, or if the SARS-Related Coronavirus 2, Isolate USA-WA1/2020, Heat Inactivated sample is subject to an issued patent.

## 6 Author Contributions

T.H.S. and H.-C.C. conceived of the project. T.H.S. performed the mathematical derivations and computational simulations. T.H.S. developed and tested the streamlined Culsans workflow and software. J.A.S. designed the IJP virus experiments. J.A.S. and T.H.S. designed the IJP aggregation experiments. J.A.S. performed all IJP experiments. J.A.S., T.H.S., and H.-C.C. analyzed and interpreted the IJP results. F.G. and T.M. designed, performed, and analyzed the AEM virus experiments. H.-C.C. and S.S. supervised the project and provided supporting advice. All authors contributed to the writing of the manuscript.

