## Supplementary Files for "Sub-Diffraction Stochastic Biosensing of Viruses in Untreated Plasma via Immuno-Janus Particle Agglutination and Flickering"

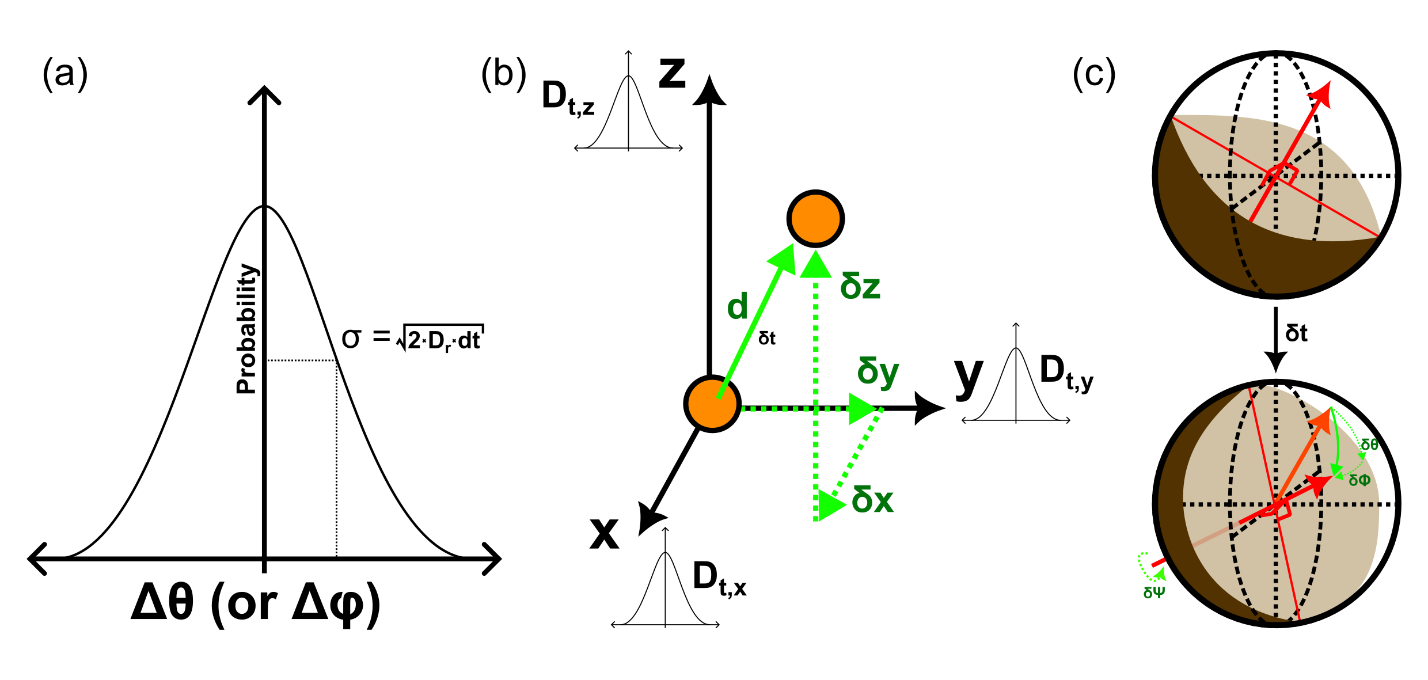

Supplementary Figure 1. Graphical representation of rotational diffusivity ($D_{r}$) for a sphere as analogously to translational diffusivity ($D_{t}$). (a) Description of rotational diffusivity as derived from the variance ($\sigma^{2}$) of a Gaussian distribution of potential rotational perturbations along each linearly independent rotational degree of freedom in a sphere $\left( \theta,\phi\right)$. This distribution is temporally self-similar by the scaling time step ($\delta t$). (b) Analogous representation of translational diffusivity in an ℝ^3^-space, where each linearly independent axis of motion $\left( x,y,z \right)$ obeys its own independent Gaussian distribution $\left( \delta x, \delta y,\delta z \right)$. (c) Graphical depiction of an example rotational perturbation of a Janus sphere within its three independent rotational axes $\left( \theta,\phi,\psi\right)$. A given rotation for the time step ($\delta t$) would yield independent components $\left( \delta\theta,\delta\phi,\delta\psi\right)$ where only the $\left( \delta\theta,\delta\phi\right)$ components yield discernible conformational changes due to the Janus symmetry.

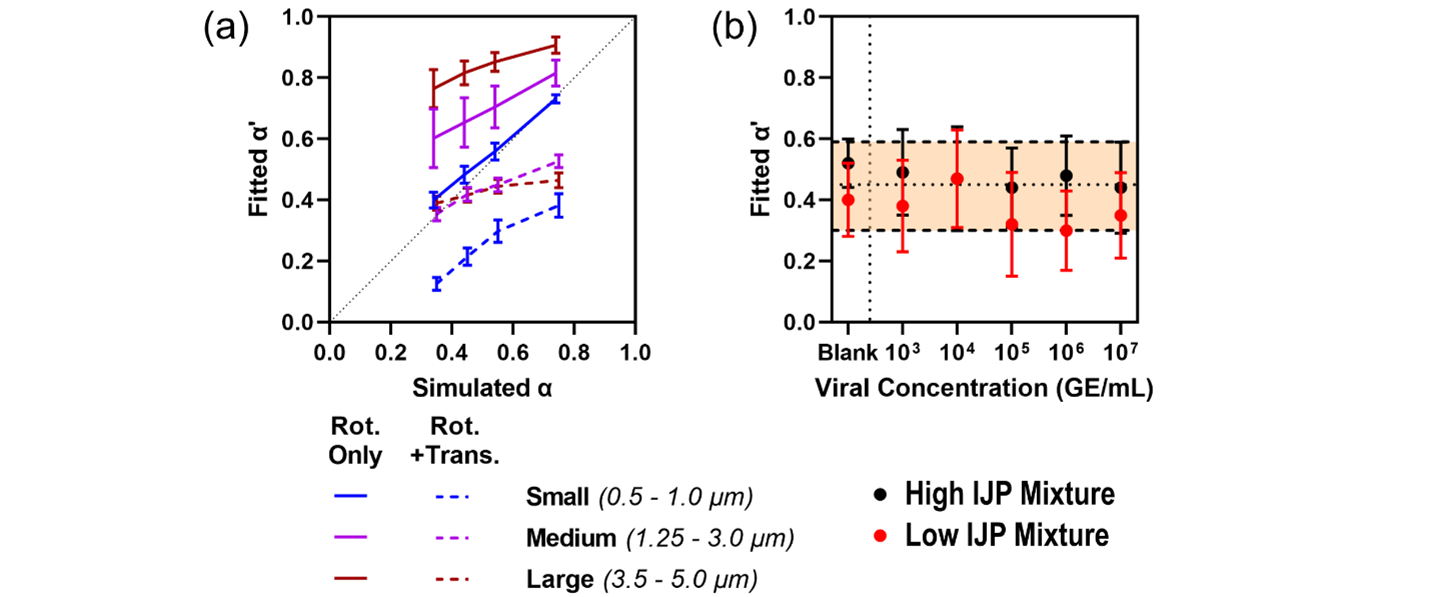

Supplementary Figure 2. Robustness testing of estimation algorithm for the fractional brightness factor $\alpha$. (a) Efficiency of approximation for the fitted $\alpha$ in recapturing simulated values of $\alpha$ for a range of particle diameters binned into small (blue series), medium (purple series), and large (red series) simulated under translationally static (Rot. Only, solid lines) and dynamic (Rot. + Trans., dashed lines) assumptions. Fitting accuracy is preferential for small particles under static conditions, whereas medium and large particles are better preserved under dynamic conditions. (b) Fitted $\alpha$-values derived from experimental time series of IJPs incubated in varying concentrations of viruses. Experiments run under standard IJP concentrations (black) and reduced IJP concentrations (red) are shown to exhibit comparable $\alpha$-values across all experiments. All error bars represent standard deviations.

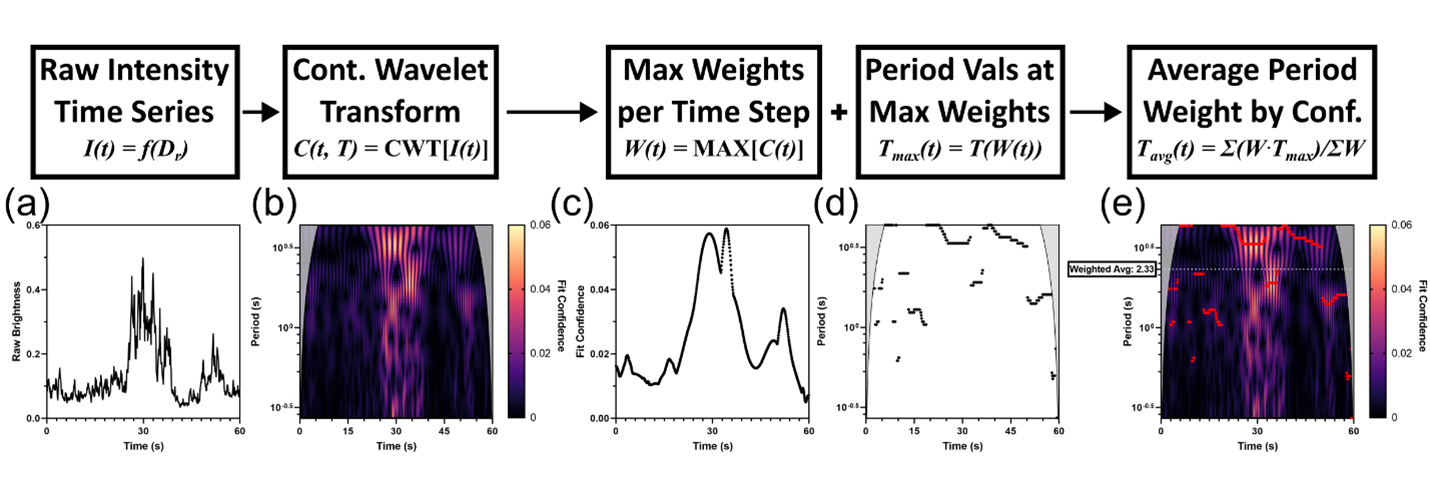

Supplementary Figure 3. Workflow of the legacy Continuous Wavelet Transform (CWT) particle track analytical technique with (a) demonstrative particle track to show analysis path. (b) The raw intensity is run through the MATLAB CWT function to produce the resultant magnitude scalogram. To adjust for edge effect interference at the start and end of the video, a Cone of Influence (gray) is superimposed. (c) The magnitude of the Fit Confidence and (d) the associated period of best fit is extracted for each frame throughout the video. (e) By taking the weighted average of the periods, the final metric is derived.

Supplementary Table 1. Slope, intercept, and $R^{2}$-values for linear regressions of calibration curves for IJP $D_{r}$ estimators using Culsans and CWT methods.^1^

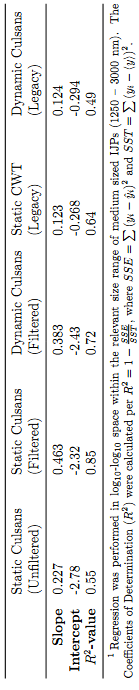

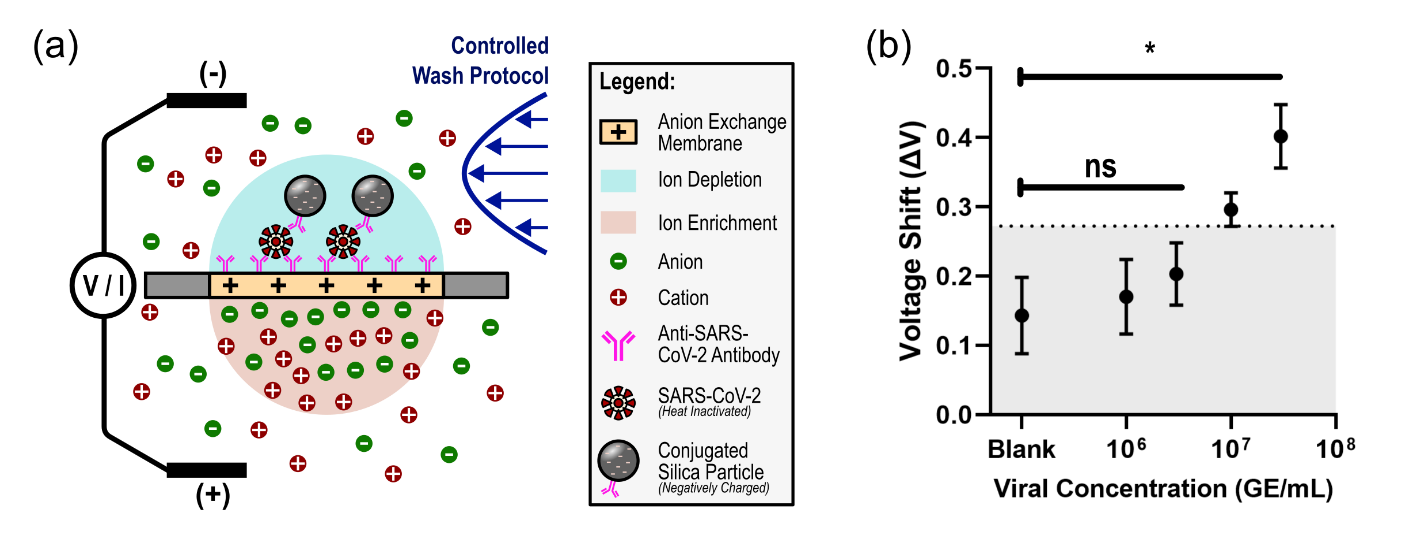

Supplementary Figure 4. An Anion Exchange Membrane (AEM)-based biosensor developed for orthogonal detection of SARS-CoV-2 virions in biofluids. (a) Representative graphic of the SARS-CoV-2 AEM biosensor platform. The AEM excludes positive charges while the applied trans-membrane electric field shuttles negatively charged anions across. This behavior produces ion-depleted and ion-enriched regions on opposing sides of the membrane which yields non-Ohmic behavior (deemed the Limiting Region) when measured using Cyclic Voltammetry. As voltages increase, local instabilities lead to electroconvection restoring Ohmic behavior (deemed the Overlimiting Region). Target antibodies are preemptively functionalized onto the ion-depletion side where biologic samples are then incubated, treated with negatively-charged reporter particles, and washed. The affixed reporter particles locally gate instability. With higher reporter concentrations, higher voltages are required to trigger Overlimiting behavior. By measuring the shift in Onset Voltage, we correlate a $log_{10}$-scale dependency of reporter concentrations to the shift in voltage signal. (b) Calibration curve generated for the SARS-CoV-2 AEM platform using serial dilutions of heat-inactivated virions spiked into healthy human plasma. Limit of detection corroborates the previously reported sub-picomolar limit at approximately ${10}^{6}$ - ${10}^{7}$ virions/mL. Each experiment was run in triplicate, and error bars represent standard deviations.

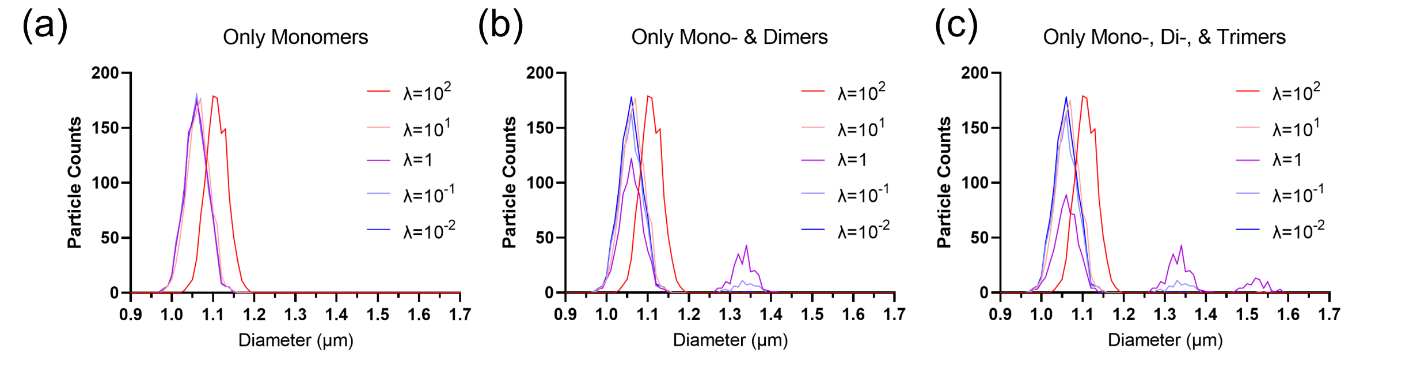

Supplementary Figure 5. Effective size distributions of numeric population simulations featuring IJPs (1 μm) binding to Poisson distributions of target nanoparticles (200 nm) and experiencing various degrees of target-bridging IJP aggregation. (a) The case of no bridging events serving as the case of the classical expected behavior where the population mean only shifts significantly for $\lambda>10$. (b) The case of bridged dimer events occurring for all sites that are assigned exactly one binding event. (c) The inclusion of an additional mechanism for a trimerization between groups experiencing exactly one and two binding events. For the cases where aggregation is allowed, the population mean now begins shifting for smaller $\lambda>0.1$.

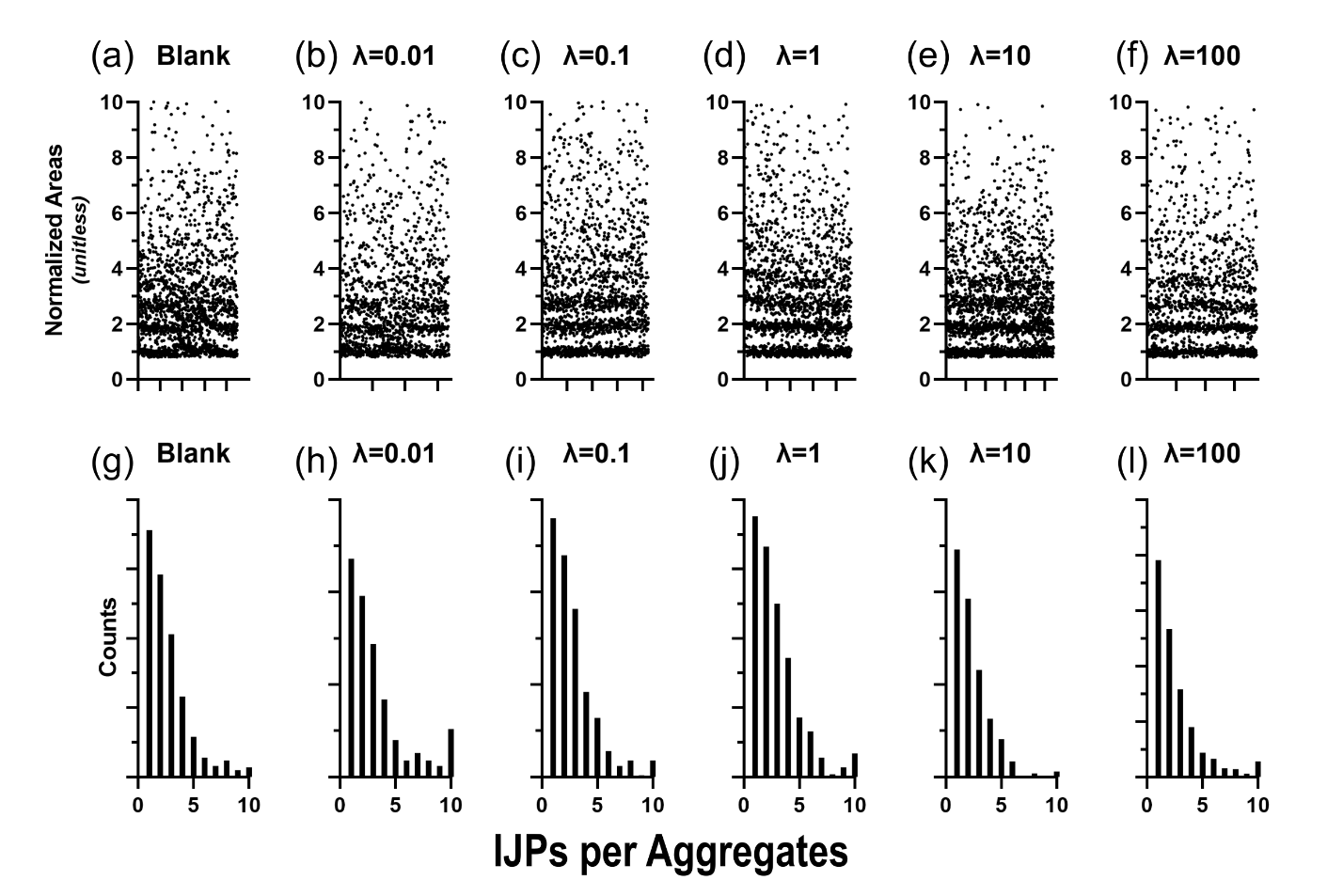

Supplementary Figure 6. Particle binning from scanning electron microscopy (SEM) images of Streptavidin-functionalized IJPs incubated with various concentrations of 200 nm biotinylated nanobeads. (a-f) Cluster areas yielded from automated particle detection from broad-field images normalized by area of IJP monomer measured for each image. Each subfigure contains particles detected from three independent images with each image containing 500 – 750 detected clusters. (g-l) Manually binned aggregate sizes counted for each image. Each subfigure contains particles detected from ten independent images with each image containing 100 - 200 separate aggregates.

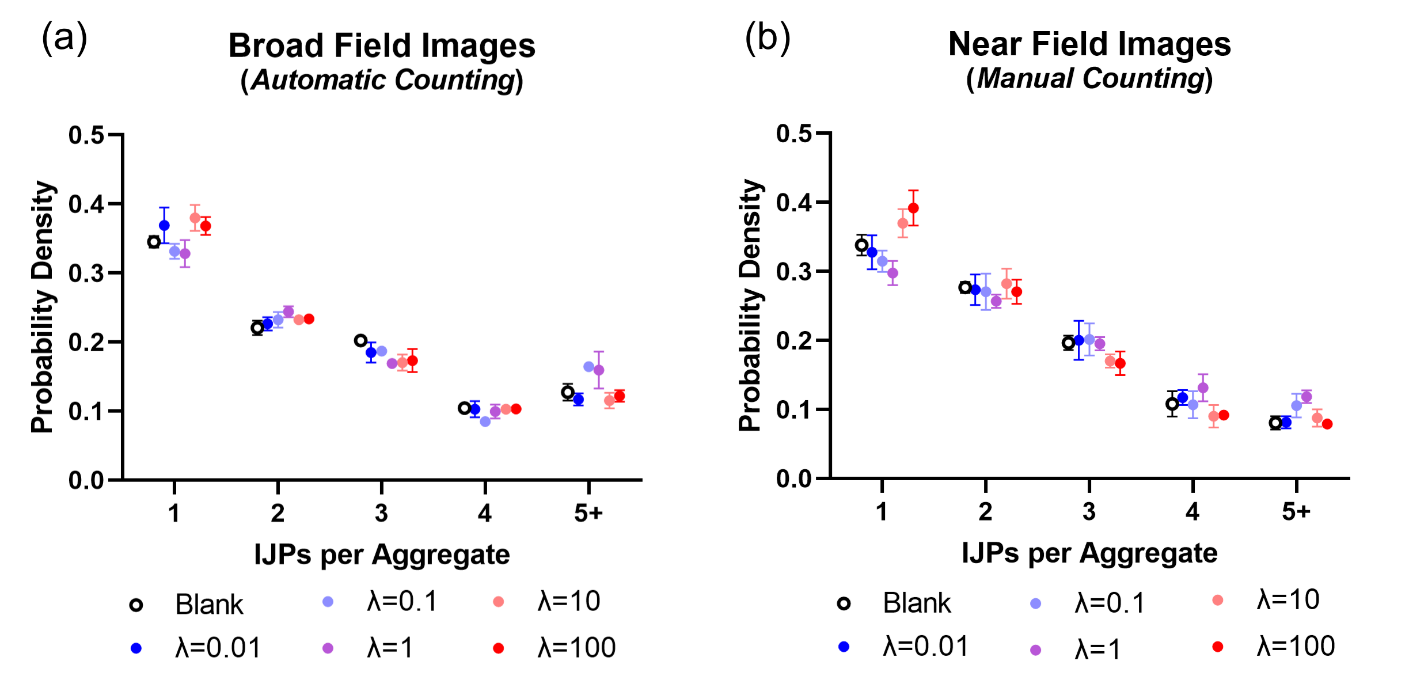

Supplementary Figure 7. Aggregate size binning based on number of IJP subunits. (a) Aggregate size percentages normalized to total number of detected aggregates derived from broad-field images using semi-automated particle detection and area mapping. Each data point represents results of $n = 3$ broad-field images. (b) Aggregate size percentages normalized to total number of detected aggregates derived from near-field images using manual counting. Each data point represents the results of $n=10$ near-field images. All error bars represent standard errors of the mean.

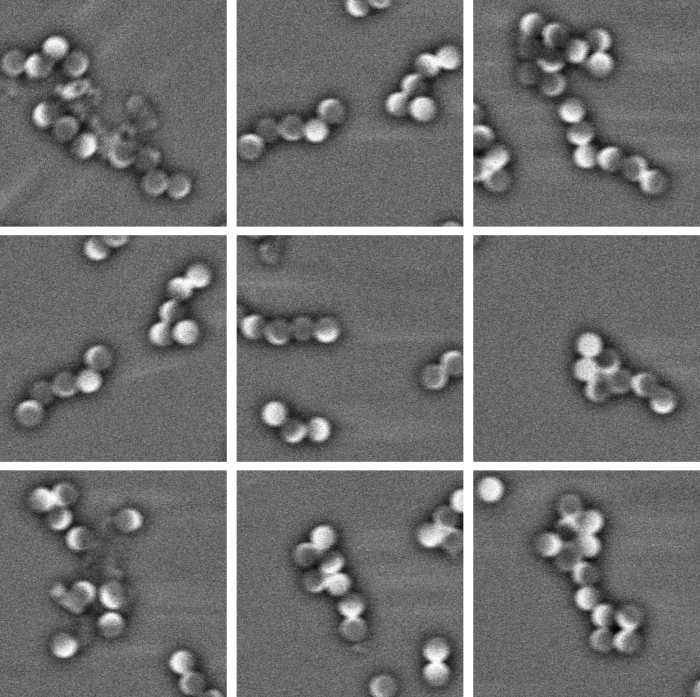

Supplementary Figure 8. Examples of chained aggregates observed in near-field images of dropcast IJPs without nanobead incubation.

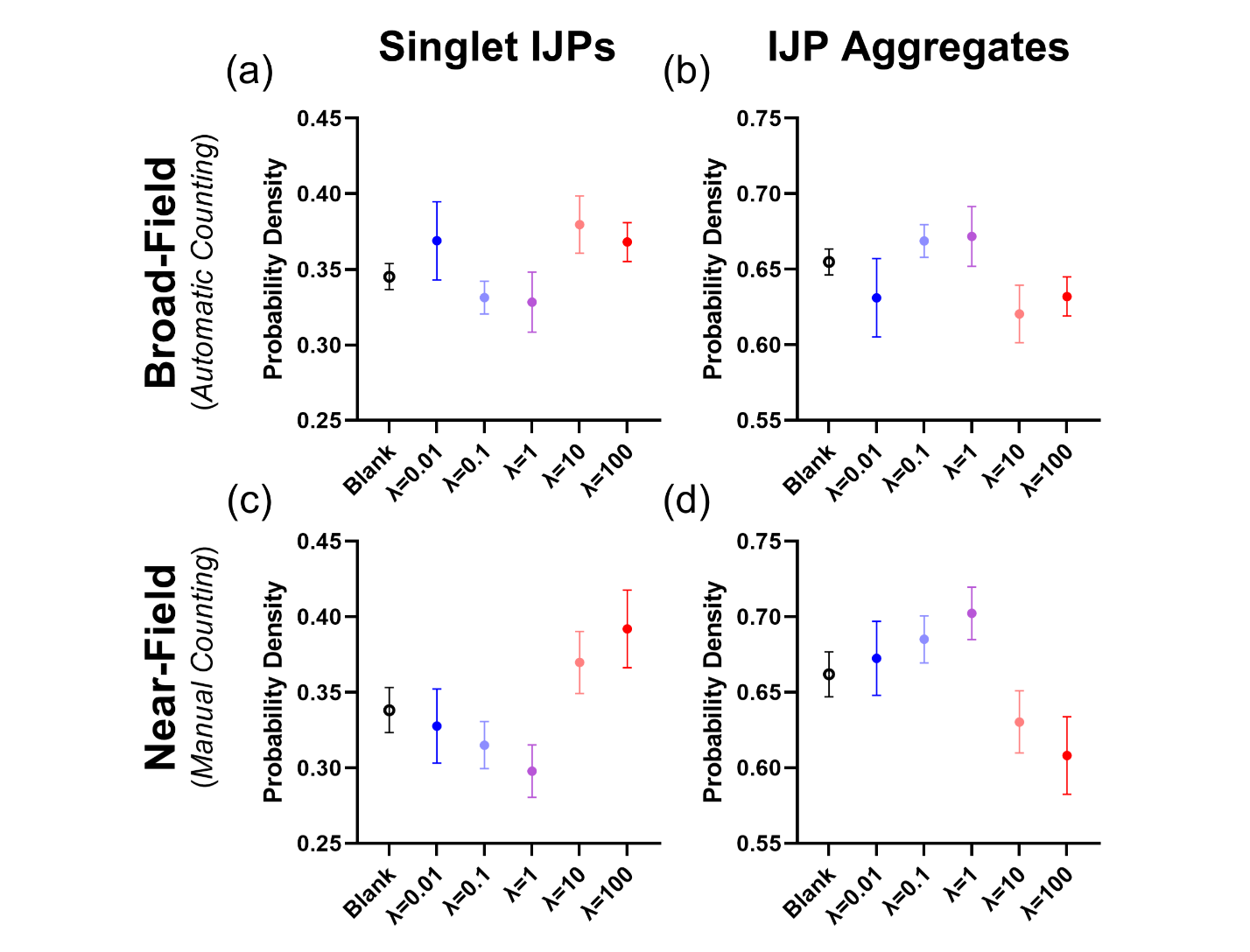

Supplementary Figure 9. Relative prevalence of IJP singlets and aggregates for varying ratios of 200 nm biotinylated nanobeads to 1 μm streptavidin-functionalized IJPs incubated together, dropcast onto glass slides, and air dried. (a - b) Population results of broad-field SEM images derived using automated particle detection. Each data point represents singlet and aggregate counts from three independent images. (c - d) Population results of near-field SEM images derived using manual aggregate counting and binning. Each data point represents singlet and aggregate counts from ten independent images. (a, c) Fraction of singlet IJPs for each condition. (b, d) Fraction of IJP aggregates composed of two to six individual IJPs. Notice the reduction in singlets and gain in aggregates with increasing $\lambda\to1$, and subsequent increase in relative IJP singlets and decrease in IJP aggregates as $\lambda\gg1$. Agglutination is most pronounced at $\lambda=1$. Error bars represent the standard errors of the mean between images.
